# Current *Schistosoma mansoni* exposure and infection have distinct determinants: a data-driven population-based study in rural Uganda

**DOI:** 10.1101/2024.04.24.24306227

**Authors:** Fabian Reitzug, Narcis B. Kabatereine, Anatol M. Byaruhanga, Fred Besigye, Betty Nabatte, Goylette F. Chami

**Affiliations:** Big Data Institute, Nuffield Department of Population Health, University of Oxford, Oxford, United Kingdom; Division of Vector-Borne and Neglected Tropical Diseases, Uganda Ministry of Health, Kampala, Uganda

**Keywords:** Exposure, water contact, schistosomiasis, *Schistosoma mansoni*, transmission, helminth, precision mapping, WASH, human behaviour, environment, ecology, snail, sub-Saharan Africa, age, gender, SchistoTrack

## Abstract

Exposure to parasitic flatworms causing schistosomiasis is a complex set of human-environment interactions. Yet, exposure often is equated to current infection. Here we studied risk factors and population patterns of exposure (water contact) within the SchistoTrack Cohort for 2867 individuals aged 5-90 years in Eastern and Western Uganda. Households within 0.34 km of waterbodies accounted for 80% of all water contact. We found a 15-year gap between population-level peak in water contact (age 30) and infection (age 15) with practically no correlation (ρ=0.03) of individual-level water contact and current infection. Bayesian selection for 30 biosocial variables was used to separately predict water contact and current infection. Water contact was positively associated with older age, female gender, fishing occupation, lack of site contamination, unsafe village drinking water, number of sites and type (beach/pond), lower village-level infection prevalence, and fewer village roads. Among these variables, only older age and fishing were positively, though inconsistently associated with infection status/intensity. Water, sanitation, and hygiene influenced water contact but not infection. Our findings highlight that exposure was highly focal and at-risk groups for exposure and infection were different. Precision mapping and targeted treatment/interventions directly focused on exposure are needed to save medicines and reduce transmission.

## Background

Environmentally mediated pathogens primarily affect populations in tropical low and middle-income countries (LMICs) and are estimated to cause an annual loss of 129,488 million disability-adjusted life years (DALYs) or approximately 40% of the global infectious disease burden [1]. Within LMICs, disease burden is concentrated in rural poor communities. Interactions of socio-demographic and environmental factors with pathogen exposure and infection acquisition have made it difficult to isolate determinants of exposure. Here we focus on schistosomes, parasitic flatworms that infect over ∼200 million people globally [2]. Schistosome transmission is complex and driven by human behaviour, accessibility of safe water and sanitation, occupation, and ecological conditions for freshwater snails that are the intermediate host of the parasite. Exposure to schistosomes occurs during water contact with lakes, rivers, or streams through activities including swimming, bathing, or fetching drinking water [3]. During water contact, cercariae—the free-living stage of the parasite—enter a human host by burrowing through the skin. With no available vaccine, mass drug administration (MDA) using praziquantel has been adopted as the main control strategy by the World Health Organization (WHO) [4]. However, treatment does not prevent reinfection and past MDA campaigns have experienced low treatment coverage, missed marginalised households, and there are concerns that repeated MDA could lead to drug resistance [5,6]. To achieve the targets set out in the 2030 WHO roadmap for neglected tropical diseases [7], there is a need to complement MDA with additional control interventions such as water access, sanitation and hygiene (WASH) provision, environmental control/modification or behaviour change [8–12]. Yet, knowledge required for identification of high exposure groups and for targeting of risk factors that determine exposure is currently limited.

Water contact has become a well-established proxy indicator for exposure due to the difficulties in directly measuring cercarial exposure. Previous studies have largely been cross-sectional and used self-reported water contact activities or constructed crude binary indicators of water contact for the purposes of predicting current infection [3]. Among 101 studies in a recent systematic review and meta-analysis by Reitzug et al., only 21.8% (22/101) collected snail abundance data to account for environmental factors relevant for translating water contact into parasite acquisition risk [3]. Attempts to integrate water contact with environmental variables have not consistently improved the ability to predict infection [13–16]. This may be due to limitations in data such as rough estimates of bodily immersion data [13,15], assumptions made about cercarial density [13,16], or methodological limitations including no systematic variable selection and no out-of-sample validation [3]. Due to regular MDA, studying the role of exposure for infection is further complicated by immunity which can be acquired through past infection or successful treatment with praziquantel [17–19]. Understanding exposure also has proven difficult as no standardised exposure measurement tools exist and there is high heterogeneity across existing studies [3].

We are lacking comparative studies on risk factors for exposure versus current infection. It has been found that infection prevalence varies based on household distance to waterbodies [20,21] but whether this is explained by corresponding trends in exposure over distance remains unclear. For estimating the force-of-infection, current mathematical models assume age-specific prevalence to be proportional to age-specific trends in current water contact [22] and do not always account for individuals who remain infected for years without current exposure due to the long lifespan of the schistosome parasite [23]. Applied statistical models of exposure have predominantly focused on exposure as a predictor of infection without characterising exposure as its own outcome and identifying shared risk factors with current infection [3].

We comprehensively characterised water contact using applied statistical models. Data was collected from January to February 2022 on the River Nile, Lake Albert, and Lake Victoria in rural Uganda as part of a population-based study within the baseline of the SchistoTrack Cohort [24]. Across 38 fishing villages in Pakwach, Buliisa and Mayuge Districts, we surveyed 2867 individuals from 1444 randomly sampled households. Socio-demographics, biomedical information, WASH information, environmental data, and water contact data were collected. *Schistosoma mansoni* infection status was ascertained using Kato-Katz stool microscopy and point-of-care circulating antigen tests. Malacological data on snails as well as waypoints of water sites, households, schools, and village centres were used to capture relevant environmental and spatial variables. We answered the following questions. What are the major human-environmental determinants of water contact and at which level (individual, household, or village) are they clustered? How do exposure determinants differ from infection determinants?

## Results

### Water contact frequency, duration, type, and timing

A study overview is presented in Fig. 1. Detailed variable definitions and characteristics of the 2867 participants are presented in Tables S1-2. Our study population was characterised by a high prevalence of self-reported water contact with open freshwater bodies (46.7%, 1339/2867, 95% confidence interval (CI) 44.9-48.5%). Among participants with current water contact, the median frequency of water contact was six times per week (interquartile range (IQR) 3-11) and the median duration in hours per week was eight (IQR 3.5-17.5). Both frequency and duration exhibited overdispersion, with only few individuals engaging in high intensity water contact (Fig. S1). The three most common water contact activities among participants were getting drinking water (17.3%, 497/2867), washing clothes with soap (16.8%, 481/2867), and fishing (12.3%, 354/2867, Table S3). These common activities were done at different times of the day and had different typical durations and frequencies. Getting drinking water and washing clothes with soap were most commonly done after sunrise (6-9 am) with 32.8% (163/497) and 34.1% (164/481) of water contacts, respectively, occurring during this time. For fishing, the early evening (5-7 pm) was the most common time accounting for 24.2% (86/354) of water contacts. The median number of weekly trips was four trips for fishing and for getting drinking water (IQRs 4-7 and 4-14, respectively), and two for washing clothes with soap (IQR 2-5). Fishing had a median duration of four hours per trip (IQR 3-4 hours) while getting drinking water and washing clothes with soap had median durations of one hour (IQRs 1-1 and 1-2, respectively).

**Figure 1.**
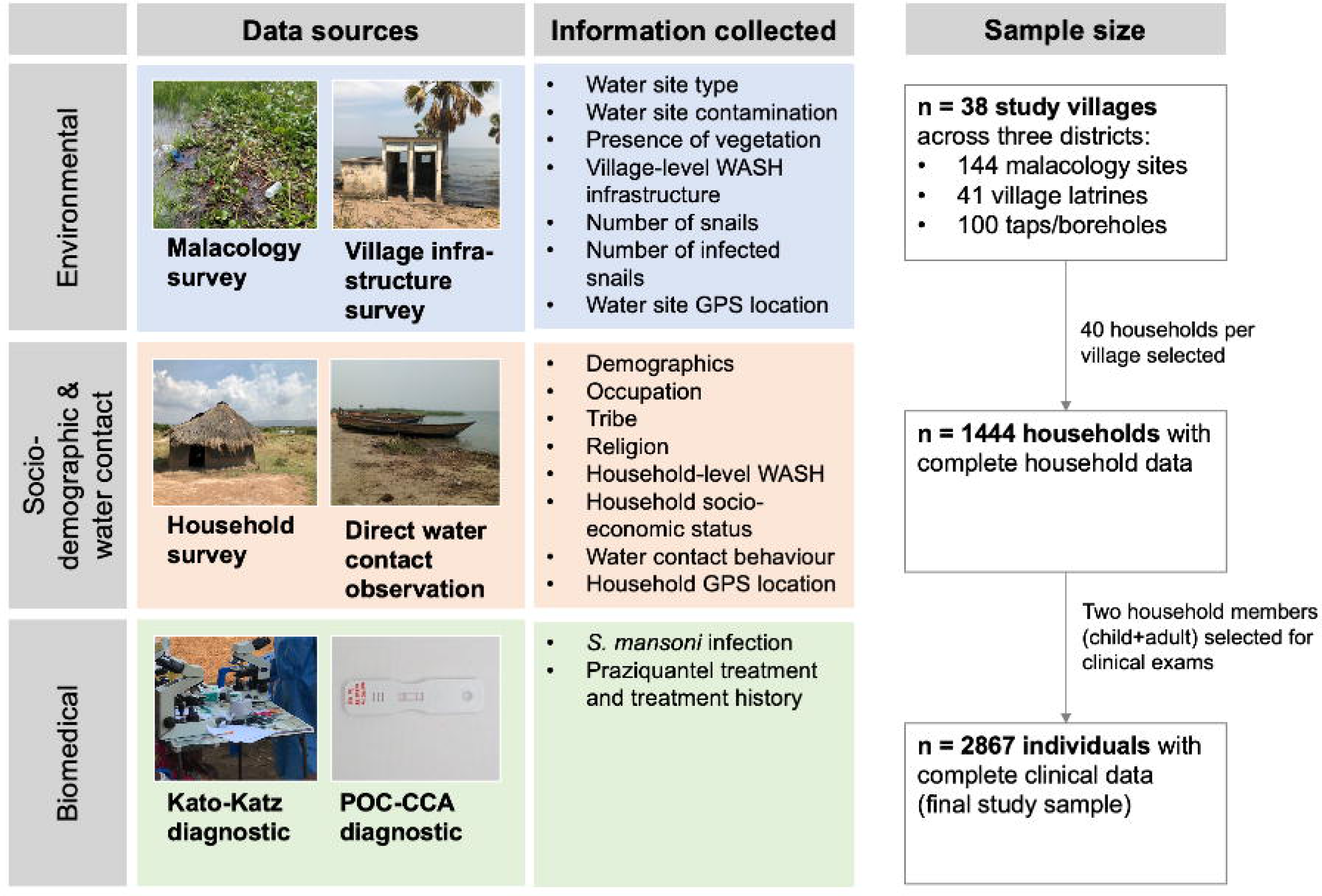
Study overview. Data sources, information collected, and sample size of the SchistoTrack study. Abbreviations: WASH = water, sanitation, and hygiene. GPS = global positioning system.

### Water contact dependence on distance to water sites

Households were located within 0.01-2.17 km (mean 0.35 km, 95% confidence interval (CI) 0.34-0.36 km) of the River Nile, Lake Albert or Lake Victoria (Fig. 2b-d). Household distance varied by district. In Pakwach District, the mean distance to the River Nile or Lake Albert was 0.41 km (95% CI 0.39-0.43 km); whereas in Buliisa District, the mean distance to Lake Albert was 0.24 km (95% CI 0.23-0.26 km). Distances in Mayuge and Pakwach were more similar even though they were the districts furthest apart. Water contact was highly concentrated around waterbodies as 80% of all participants with water contact (and 64.3% of all participants) lived in households within 0.34 km Euclidean distance of a water site (Fig S2). In households residing directly on the shoreline (≤100 metres), 54% of individuals (315/589) had water contact. In households located >1 km from a water site, 26% (36/137) reported having water contact. We used generalised additive models (GAMs) to examine the influence of household distance to the closest water site on the prevalence of water contact. For every 100m increase in distance between 0-1 km, we found a 3.4% absolute reduction in the water contact. Consistently decreasing gradients were observed across domestic, occupational, and recreational activities (Fig. 3a). Gradients in water contact differed depending on the distance metric used (Figs. 3b-d). Village centre distance to the closest water site produced somewhat weaker gradients than household distance (1.9% absolute reduction for every 100m-increase with village distance versus 3.4% with household distance, Fig. 3c). When using distance of primary schools to the closest freshwater body there was no relationship between distance and the percentage of households who engaged in water contact (Fig. 3d).

**Figure 2.**
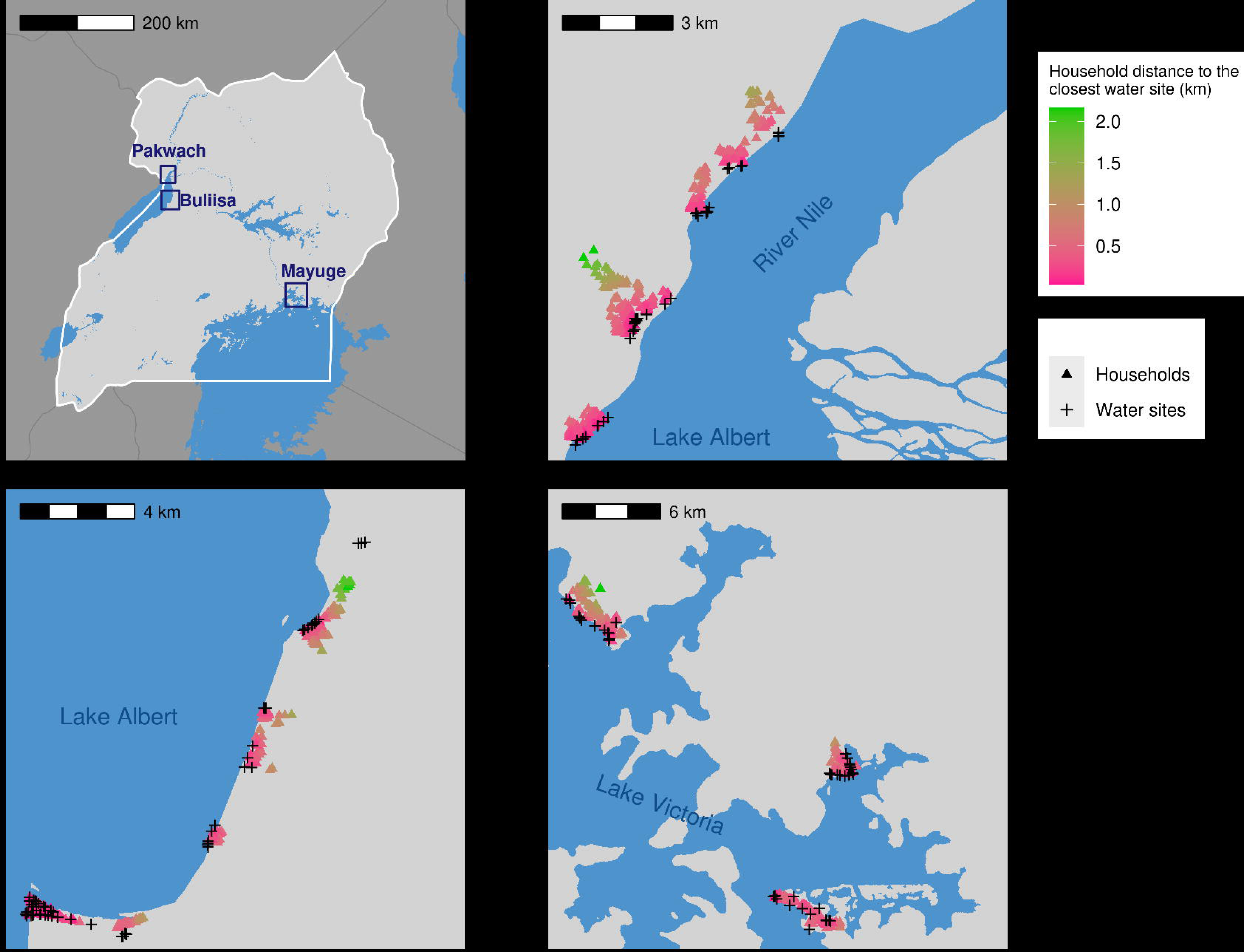
Study areas. Global positioning system (GPS) locations of 2867 study households (randomly displaced by 50m) and 144 water sites sampled by malacologists. Waterbody boundaries from the World Resources Institute. **A**. Locations of the three study districts Pakwach, Buliisa, and Mayuge in Uganda. **B**. Detail of Pakwach: location of study households and water sites along Lake Albert and the River Nile. **C.** Detail of Buliisa: location of study households and water sites along Lake Albert. **D.** Detail of Mayuge: location of study households and water sites along Lake Victoria. Household distance in this figure represents Euclidean distance of the household location to the closest water site.

**Figure 3.**
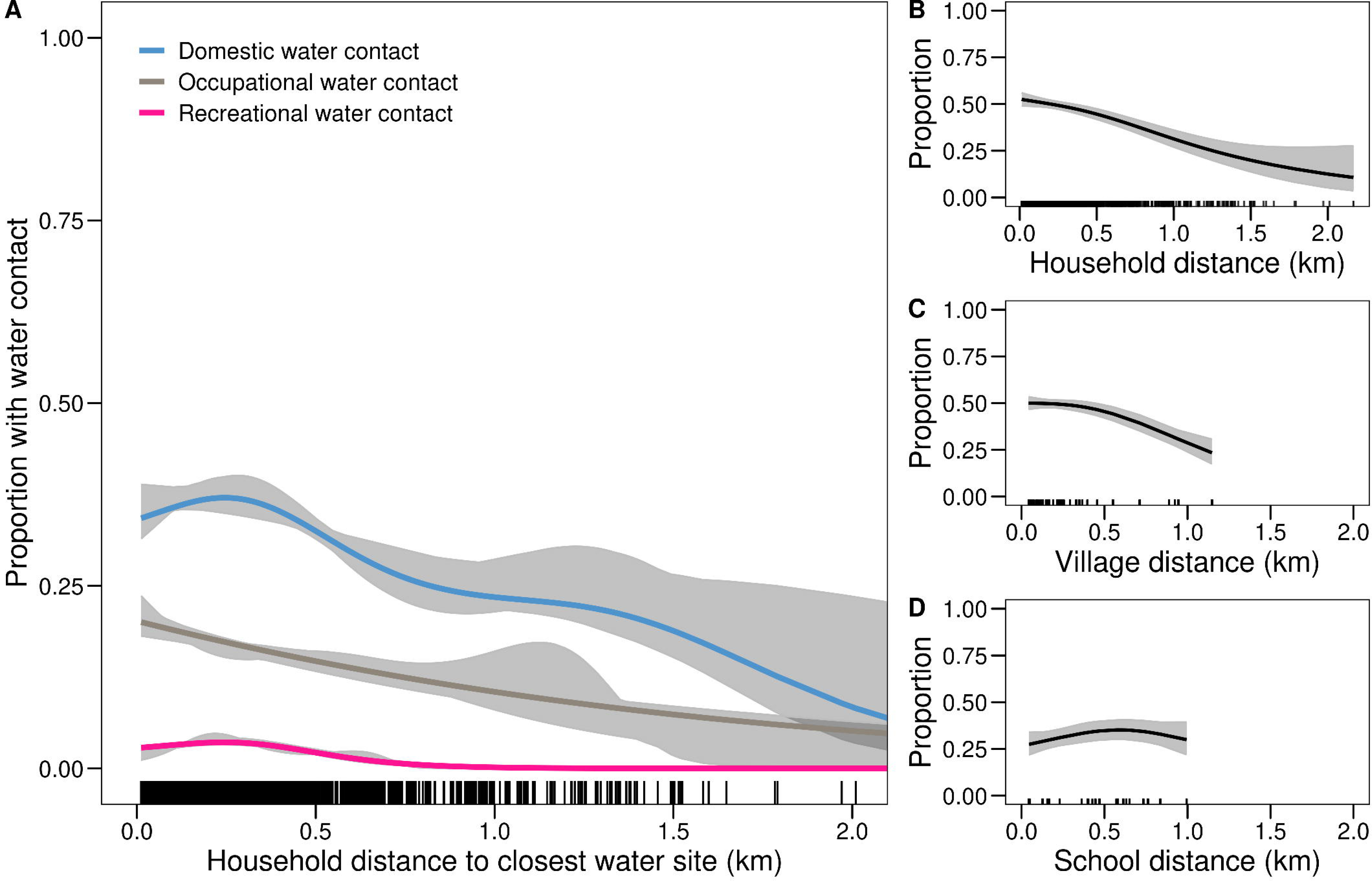
Variation in the prevalence of water contact over waterbody distance. Proportion of participants having current water contact over Euclidean household distance to the closest water site (in km) modelled using generalised additive models. **A**. Proportion of participants with domestic, occupational, and recreational water contact. **B-D**. Comparison of lake gradients in water contact across alternative waterbody-distance metrics: household distance, village centre distance, and school distance to the closest water site. **B.** Proportion of participants with any water contact by household-level distance to the closest water site. **C**. Proportion of participants with water contact by village centre distance to the closest water site. (Village centre location as reported by the village chairman. All households within a village were assigned the distance of the village centre to the closest water site.) Compared to household water site distance with a maximum of 2.2 km, the village centre distance distribution was truncated at 1.2 km as the furthest village centre in this study was 1.2 km away from the closest water site. **D**. Prevalence of water contact by distance from the school closest to the household to the nearest water site. Sample restricted to n=597 enrolled children. The 95% confidence intervals in Panel A were computed via bootstrap with 1,000 repeats.

### Gender and age-specific water contact patterns

We used GAMs to examine gender- and age-specific patterns of water contact. Overall, 43.5% (775/1573) of females and 49.3% (564/1294) of males reported water contact. The duration of water contact was higher for females than males (median of 7 vs 5 trips per week, respectively, Wilcoxon rank sum test p<0.01, Table S4). Females had a lower duration of water contact than males (7 vs 10 hours per week, respectively, Wilcoxon rank sum test p<0.01, Table S5). Relationships between current water contact with age and gender were nonlinear and varied substantially over the life course. Modelled age-dependent prevalence of water contact was 18% (95% CI 14-18%) at age 5, peaked at 70% (95% CI 66-74%) at age 30, and declined to 28% (95% CI 20-36%) at age 70 (see Fig. 4a). Females had a lower prevalence of water contact than males across ages 18-35 (Fig. S3). Water contact activities were gender-dependent (Table S3). For instance, 24.3% (315/1294) of males reported going fishing while the same figure was only 2.5% (39/1573) among females (χ² = 311.56, p<0.01). Collecting drinking water was reported by 22.1% (348/1573) of females but only 11.5% (149/1294) of males (χ² = 55.02, p<0.01). Washing clothes with soap was also more prevalent among females than males (23.4% (368/1573) versus 8.7% (113/1294), respectively, χ² = 108.27, p<0.01). The proportion of water contacts taking place during peak cercarial shedding time (10am-3 pm) was not significantly different between males and females for any activity. However, predominantly female activities (getting drinking water, washing clothes with soap, washing jerry cans or household items) were more likely to be conducted during peak shedding time than male activities such as fishing (Table S6). Gender differences in water contact frequency, duration and dominant activities became pronounced between ages 15-19 (Fig. 4b-c). Until age 15, water contact patterns of both females and males included mostly domestic activities. After age 15, females maintained high levels of domestic water contact while the relative involvement of males in domestic activities strongly declined. Among adults (age 18+), domestic water contact accounted for 75.3% of total water contact duration for females while occupational water contact accounted for 81.8% of total water contact duration for males (Fig. 5).

**Figure 4.**
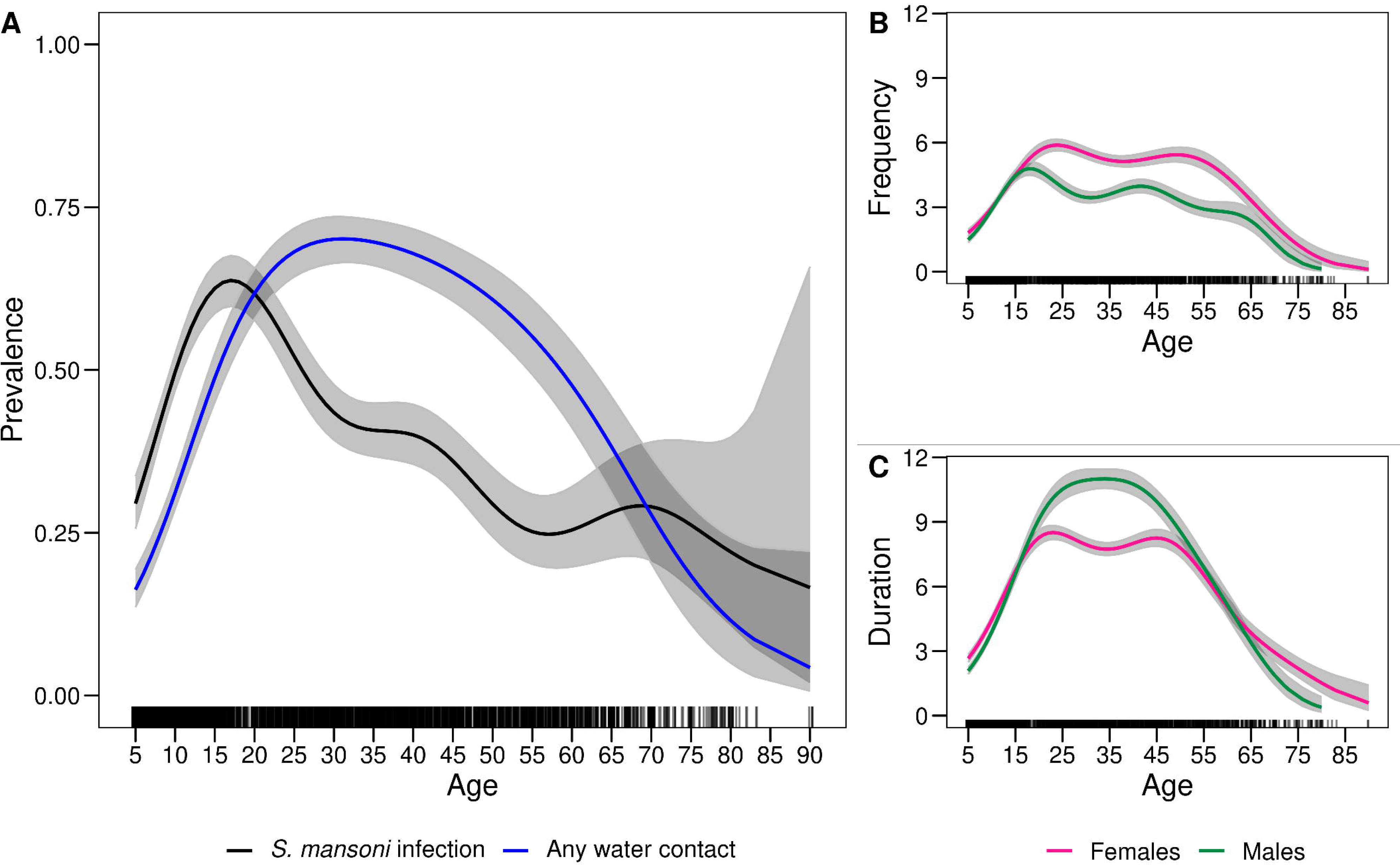
Variation in water contact and infection over age. **A.** Proportion of participants with current water contact and *S. mansoni* infection prevalence over age modelled using generalised additive models. **B**. Frequency of water contact measured by the weekly number of trips to waterbodies over age and gender. **C**. Duration of water contact measured in numbers of hours per week, over age and gender.

**Figure 5.**
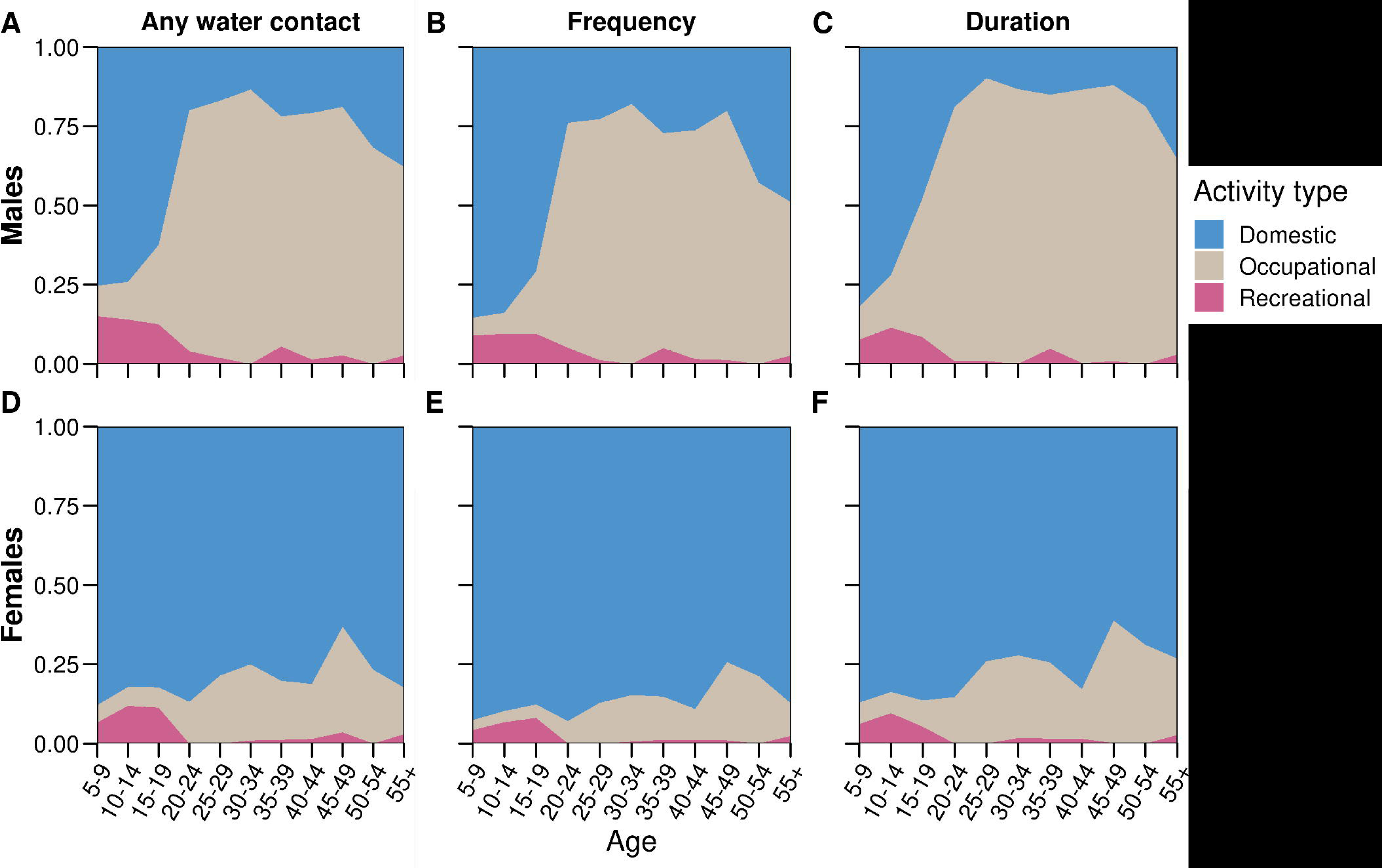
Composition of water contact activities over age and gender. **A, D.** Proportion of all water contacts within each 5-year age group which were domestic, occupational, or recreational over age and gender. **B, E.** Frequency, measured as the proportion of trips to waterbodies which were domestic, occupational, or domestic over age and gender. **C, F.** Duration, measured as the proportion of water contact time which was accounted for by domestic, occupational, or recreational activities over age and gender.

### Human-environmental determinants of water contact

To identify human-environmental determinants of water contact, we used Bayesian variable selection (BVS) on a comprehensive set of 30 variables (ten socio-demographic, one biomedical, ten WASH, and nine environmental candidate variables, see Fig. 6a-d). Variables with inclusion probabilities ≥0.5 were used in multivariable logistic regression models to predict water contact. Selected socio-demographic factors were age, age^2^, gender, and occupation. One village-level WASH variable –the proportion of households using a safe drinking water source per village— was included, as well as village infection prevalence. Four environmental variables were selected: type of water site closest to household, number of water sites per village, roads per village, and site contamination occurring at the closest site to the household. To summarise which variable types and levels were relevant for predicting water contact, we calculated the proportion of variables by group and level, as weighted by their respective inclusion probabilities (Fig. 6e-f). Among selected variables, socio-demographics accounted for 43% of all weights, followed by environmental variables (35%), WASH (16%), and biomedical variables (6%, Fig. 6e). Individual-level factors accounted for 30%, household-level factors for 41%, and village-level factors for 29% of weights, respectively (Fig. 6f).

**Figure 6.**
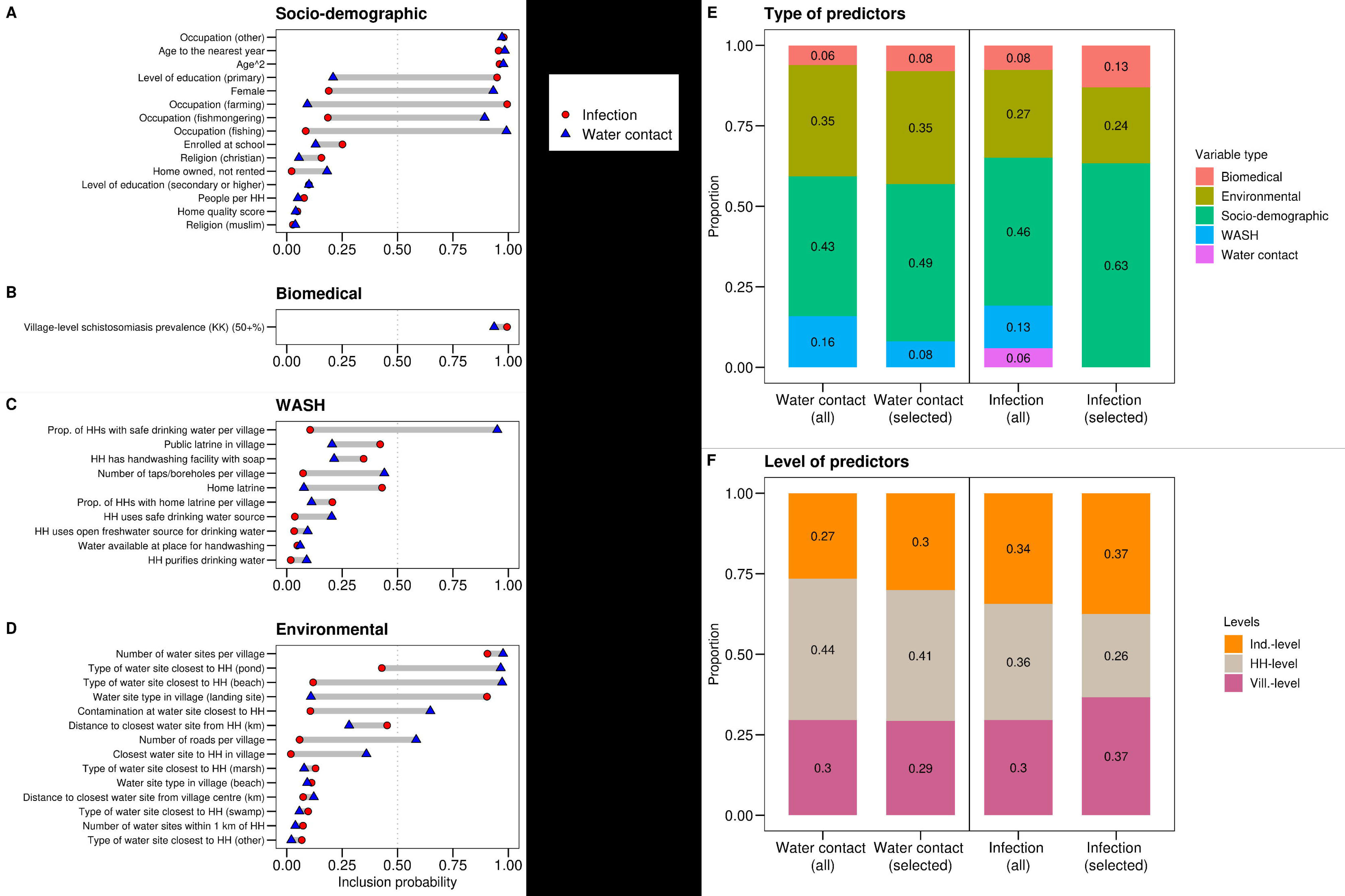
Relevant predictors of exposure and infection. **A-D.** Relevance of socio-demographic, biomedical, WASH, and environmental variables for predicting water contact and infection, as indicated by their inclusion probability in the ‘best predictive model’ (median probability model) based on Bayesian variable selection (BVS). The dashed vertical line represents the cut-off (pr ≥ 0.5) used to select variables for inclusion in the final regression models. **E-F** regroup variables in A-D according to their type and level and show the importance of different levels and types of variables as indicated by their proportional representation in the variable set (weighted by their respective inclusion probabilities). Within each box in E-F, the left column shows the proportion each variable category accounts for within the candidate variable set and the right column shows the importance of each category within the set of selected variables. Abbreviations: Prop. = proportion. HH = household. WASH = water, sanitation, and hygiene. Ind.-level = individual-level. Vill.-level = village-level.

Fig. 7 presents the main exposure model. Significant predictors of current water contact spanned socio-demographic, biomedical, WASH, and environmental variables. A one-year increase in age was associated with a 1.18 times higher likelihood of having water contact (95% CI 1.15 – 1.21). The age^2^ term (odds ratio (OR) 0.9978; 95% CI 0.9975 – 0.9982), indicated significant nonlinearity but only a 0.0022% decrease with each one-year age increase. Females had 1.4 times (95% CI 1.17 – 1.68) higher likelihood of having water contact compared to males. Fishing and fish mongering occupations were associated with 6.83 times (95% CI 4.15 – 11.23), and 2.29 times (95% CI 1.22 – 4.32) higher likelihood of water contact, respectively. When contamination was observed at the water site closest to the household, people were 22% less likely to have water contact (OR 0.78, 95% CI 0.65 – 0.99). Water site ecology influenced water contact behaviour; people who lived closest to a beach or pond had a significantly higher likelihood of having water contact compared to people living closest to a river (OR beach 1.68; 95% CI 1.22 – 2.31. OR pond 2.11; 95% CI 1.35 – 3.30). The number of sites per village was associated with 1.19 times (95% CI 1.11 – 1.26) higher likelihood of water contact. In villages with ≥50% *S. mansoni* prevalence, people were 31% less likely to engage in water contact compared to villages with 10-49% prevalence (OR 0.69; 95% CI 0.58 – 0.83). Every additional road in the village was negatively associated with water contact (OR 0.84; 95% CI 0.78 – 0.91). In terms of WASH, we found that in villages where all study households used a safe drinking water source (taps/boreholes), individuals had a 65% lower likelihood of having water contact compared to people living in villages where all study households used surface water (OR 0.35, 95% CI 0.24 – 0.52).

**Figure 7.**
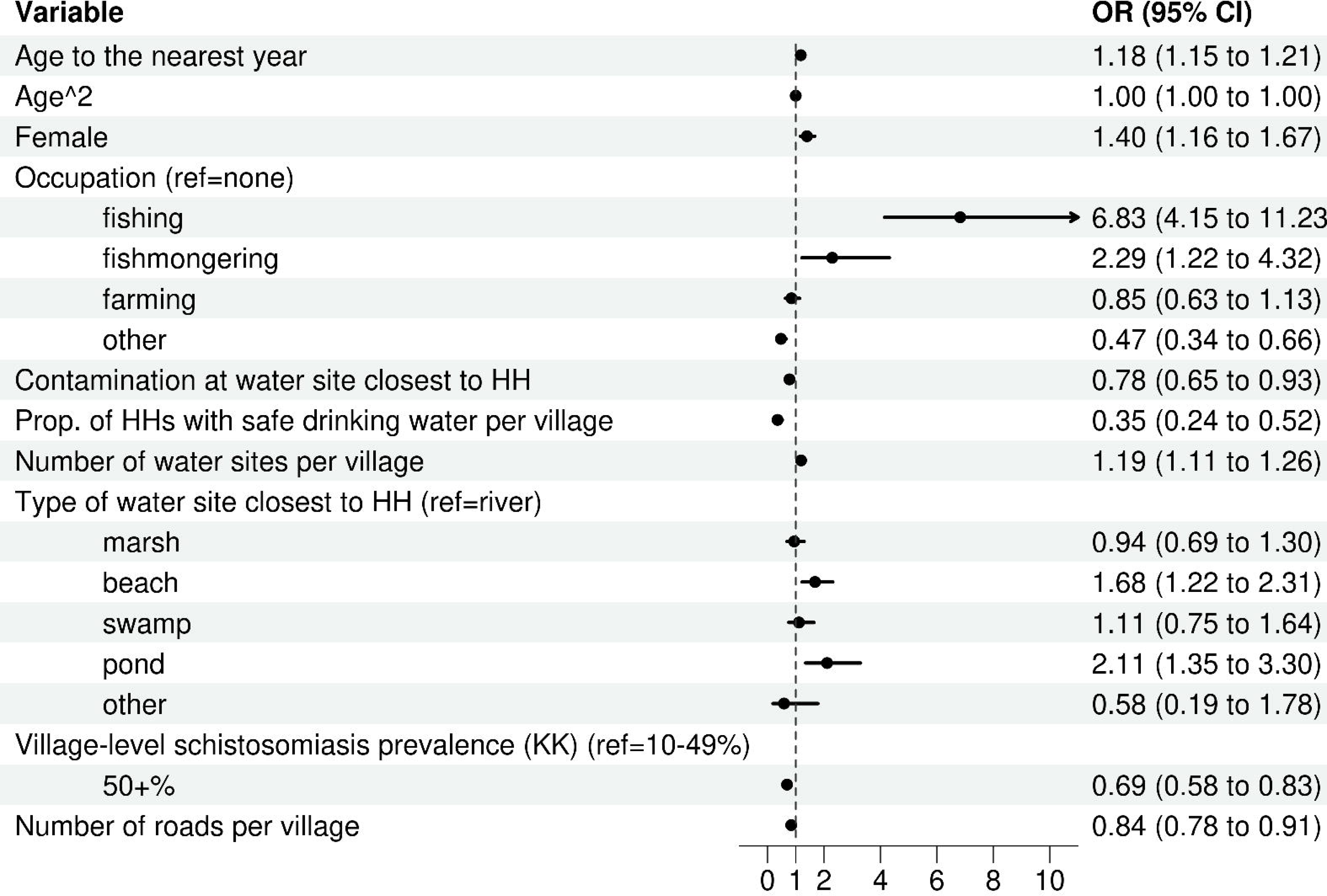
Regression model of water contact. Logistic regression model predicting current water contact (n=2867) with 95% confidence intervals from standard errors clustered at the household-level. For each categorical variable, we tested group-level significance using likelihood ratio tests (LRTs) comparing the full model to a model without the categorical variable. We found that occupation (χ^2^ = 153.4, p-value<0.01) and type of water site closest to the household (χ^2^ = 42.8, p-value<0.01) significantly improved model fit. Abbreviations: Prop. = proportion. HH = household. KK = Kato-Katz stool microscopy.

Separate logistic regression models predicting domestic, occupational, and recreational water contact showed that the effect of gender varied between activity types (Fig. S4). Females were 2.45 times more likely to have domestic water contact than males (OR 2.45; 95% CI 1.99 – 3.03), but 66% and 83% less likely to have recreational and occupation water contact (OR 0.34; 95% CI 0.13 – 0.91 and OR 0.17; 95% CI 0.11 – 0.28, respectively). Domestic water contact was 76% lower in villages where all study households used safe drinking water (OR 0.24; 95% CI 0.16 – 0.37), but no such negative relationship between safe drinking water with recreational or domestic water contact was found (OR 1.46; 95% CI 0.14 – 15.14 and OR 3.21; 95% CI 1.46 – 7.04, respectively). When restricted to individuals with water contact, negative binomial models predicting water contact frequency and duration showed no significant gender differences in duration and frequency after adjusting for covariates. Again, the proportion of study households using a safe drinking water source was associated with both lower frequency and duration of water contact (Figs. S5-6).

### Determinants of exposure vs infection

We compared determinants of exposure and schistosome infection status, as measured by Kato-Katz, to understand if risk factors of exposure and infection were distinct. Overall, 43.3% (1240/2867, 95% CI 41.4%-45.1%) of study participants were infected with *S. mansoni* (≥1 eggs per gram of stool), and 8.2% (236/2867, 95% CI 7.2%-9.2%) were heavily infected (400+ eggs per gram). District-level prevalence was highest in Pakwach (50.4%, 477/947), followed by Buliisa (44.1%, 422/958), and Mayuge (35.4%, 341/962) respectively. Modelled *S. mansoni* infection prevalence varied over age; 29% (95% CI 25-33%) of children were infected at age five, prevalence rose to a peak of 63% (95% CI 59-67%) at age 15 and then declined to 31% (95% CI 24-36%) at age 50 (Fig. 4a). This trend differed from age-dependent water contact which peaked much later (at age 30) and remained at a significantly higher level than infection until age 65. Modelled age-specific infection prevalence trends in GAMs were similar between participants with and without water contact (Fig. S7). GAMs also indicated that infection prevalence showed less of a decline over household distance to the closest water site compared to water contact (1.9% versus 3.4% average declines from every 100m increase between 0-1 km distance, respectively, Fig S8). The overall correlation between water contact and infection status was nearly non-existent (ρ=0.03, p=0.11, obs. 2867). Pairwise correlations of infection status or water contact between adults and children within the same household were weak (ρ=0.16, p<0.01, and 0.08, p=0.01, obs. 2867, respectively).

We selected predictors of infection status using the same methods and candidate set as for water contact (plus six additional water contact variables, see Table S1). The number of variables selected for water contact was ten compared to seven for infection status. Among selected predictor variables of water contact, only 5/10 (50%) were selected for inclusion in the infection status model. Shared variables were age, age^2^, occupation, number of water sites per village, and village infection prevalence. No exposure or WASH variable met the threshold of inclusion for infection status. Predictors of water contact which were not selected for infection related primarily to village-level ecology and infrastructure (type of water site closest to the household, proportion of households using safe drinking water, and roads per village). There were only two predictors of infection which were not selected for the water contact model: level of education and the type of water site in the village. When grouped by variable type, socio-demographics (accounting for 63% of variables) and environmental variables (24%) were most relevant for infection (Fig. 6e). Socio-demographics were comparatively more relevant for infection status than for water contact while environmental variables were less important for infection status compared to water contact. When compared by level, household-level variables were comparatively less relevant for infection status than for water contact while individual-level variables were more relevant for infection status than for water contact (Fig. 6f).

Fig. 8 shows results of logistic regressions predicting infection status and heavy infection. Among the five variables consistently selected for water contact and infection, no variable was significant and of similar magnitude, defined as having overlapping CIs, between the models. While age was positively associated with both outcomes, each one-year increase in age was significantly more strongly associated with likelihood of water contact (OR 1.18; 95% CI 1.15 – 1.21) than with infection (OR 1.05; 95% CI 1.03 – 1.08). We found no significant correlation between residuals of water contact and infection models (ρ=0.01, p=0.40, obs. 2867). After accounting for endogeneity of water contact and infection using bivariate logit models with all consistently selected variables, we still found associations of age and fishing with water contact and infection remained inconsistent (Table S7). Even among infection outcomes in Fig. 8, associations were inconsistent as fishing was associated with heavy infection (OR 1.74; 95% CI 1.04 – 2.91) but not with infection (OR 1.16; 95% CI 0.82 – 1.65) and having attained primary education was associated with a higher likelihood of infection but not with heavy infection (OR infection 1.53; 95% CI 1.20 – 1.94. OR heavy infection 1.09; 95% CI 0.70 – 1.68).

**Figure 8.**
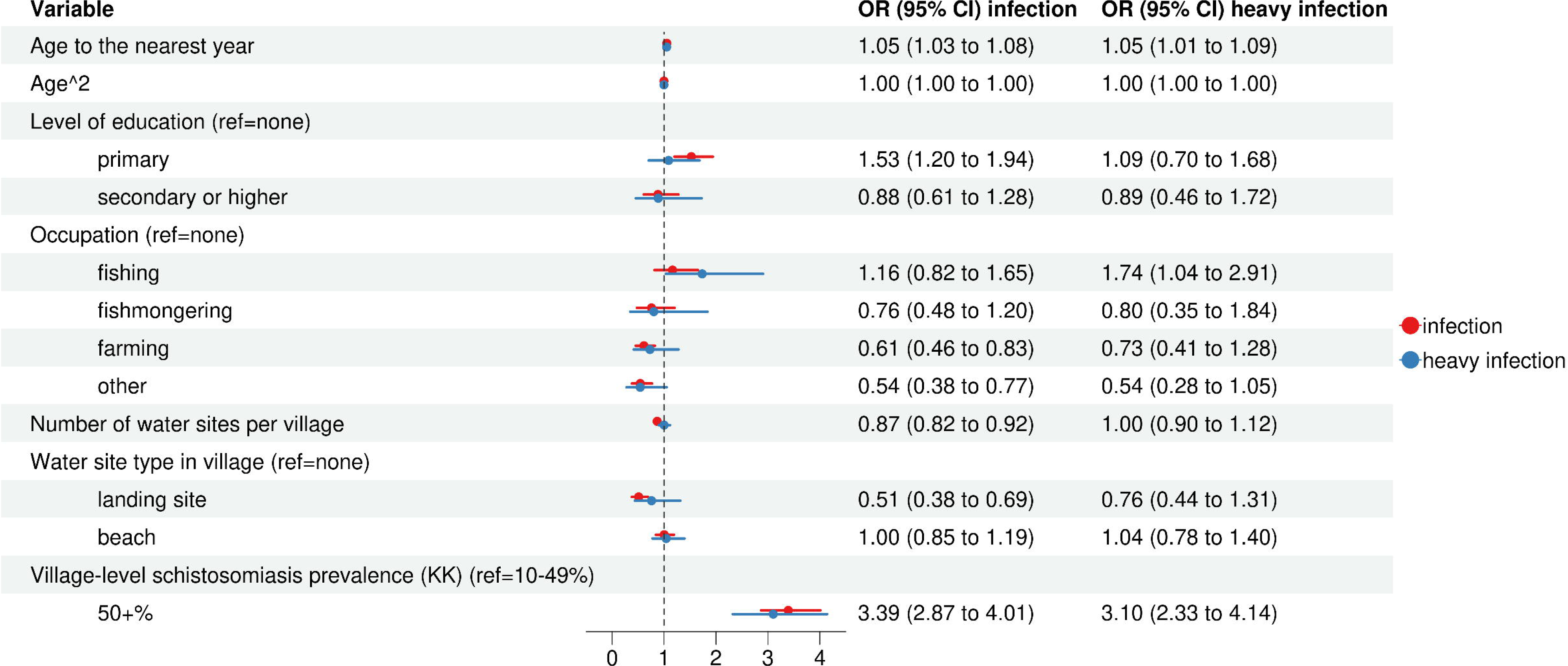
Regression models of infection status and heavy infection. Separate logistic regression model predicting *S. mansoni* infection and heavy *S. mansoni* infection (n=2867) with 95% confidence intervals from standard errors clustered at the household-level. In addition to the findings reported in the main text, we also found that village-level *S. mansoni* prevalence of ≥50% was positively associated with infection status and heavy infection intensity (OR infection 3.39; 95% CI 2.87 – 4.01. OR heavy infection 3.10; 95% CI 2.33 – 4.14) while being negatively associated with water contact (Fig. 7). School enrolment, which may approximate past school-based MDA, was not significant in any models. The number of water sites per village and having a landing site in the village were negatively associated with infection likelihood (OR 0.87; 95% CI 0.82 – 0.92 and OR 0.51; 95% CI 0.38 – 0.69, respectively) but were insignificant for heavy infection intensity. For each categorical variable, we tested group-level significance using likelihood ratio tests (LRTs) comparing the full model to a model without the categorical variable. We found that school enrolment (χ^2^ = 22.2, p-value<0.01), occupation (χ^2^ = 27.9, p-value<0.01) and type of water site in the village (χ^2^ = 19.48, p-value<0.01) significantly improved model fit in the infection status model. For the heavy infection model, only occupation significantly improved model fit (χ^2^ = 17.0, p-value<0.01) whereas school enrolment (χ^2^ = 0.65, p-value=0.72) and type of water site in the village (χ^2^ = 1.25, p-value=0.53) did not. Abbreviations: KK = Kato-Katz stool microscopy.

### Validation of exposure and infection measures

We investigated exposure misclassification in our survey data by using direct water contact observation as an alternative exposure data source. Observation data was available for 12/38 study villages and collected at the same time as the self-reported data. A comparison of the age structure observed in the direct water contact data and self-reported data suggested that the self-reported water contact data closely resembled observed community-wide water contact patterns from direct observation (Fig. S9). To address the possibility of missed light infections due to low sensitivity of Kato-Katz microscopy, we recoded negative Kato-Katz results as positive when POC-CCA diagnostic results were positive, recording trace as negative. Fig. S10 showed that infection results remained similar after reclassification of infection status.

### Model validation and relevance of snail and exposure variables for predictive accuracy

We assessed the predictive performance of the water contact and infection models using 10-fold cross-validation and quantified the influence of variable selection method (BVS versus likelihood ratio tests (LRTs)) and the relevance of including additional snail and water contact variables (Fig. 9). The water contact model had better predictive performance than the infection status model with an area under the receiver operating curve (auROC) of 0.783 versus 0.694 (p<0.01, Fig. 9a). Neither the inclusion of additional snail variables nor the inclusion of more granular water contact variables significantly improved the predictive accuracy of any model (Fig. 9b-c, see methods for details). BVS outperformed LRTs for water contact prediction (auROC 0.783 versus 0.525, respectively, p<0.01, Fig. 9c, Supplementary Fig. 10). Compared to water contact, variable sets selected for infection status were more comparable between BVS and LRTs (Table S6), and the gap in performance was smaller (auROC 0.694 and 0.625, p<0.01, Fig. 9d).

**Figure 9.**
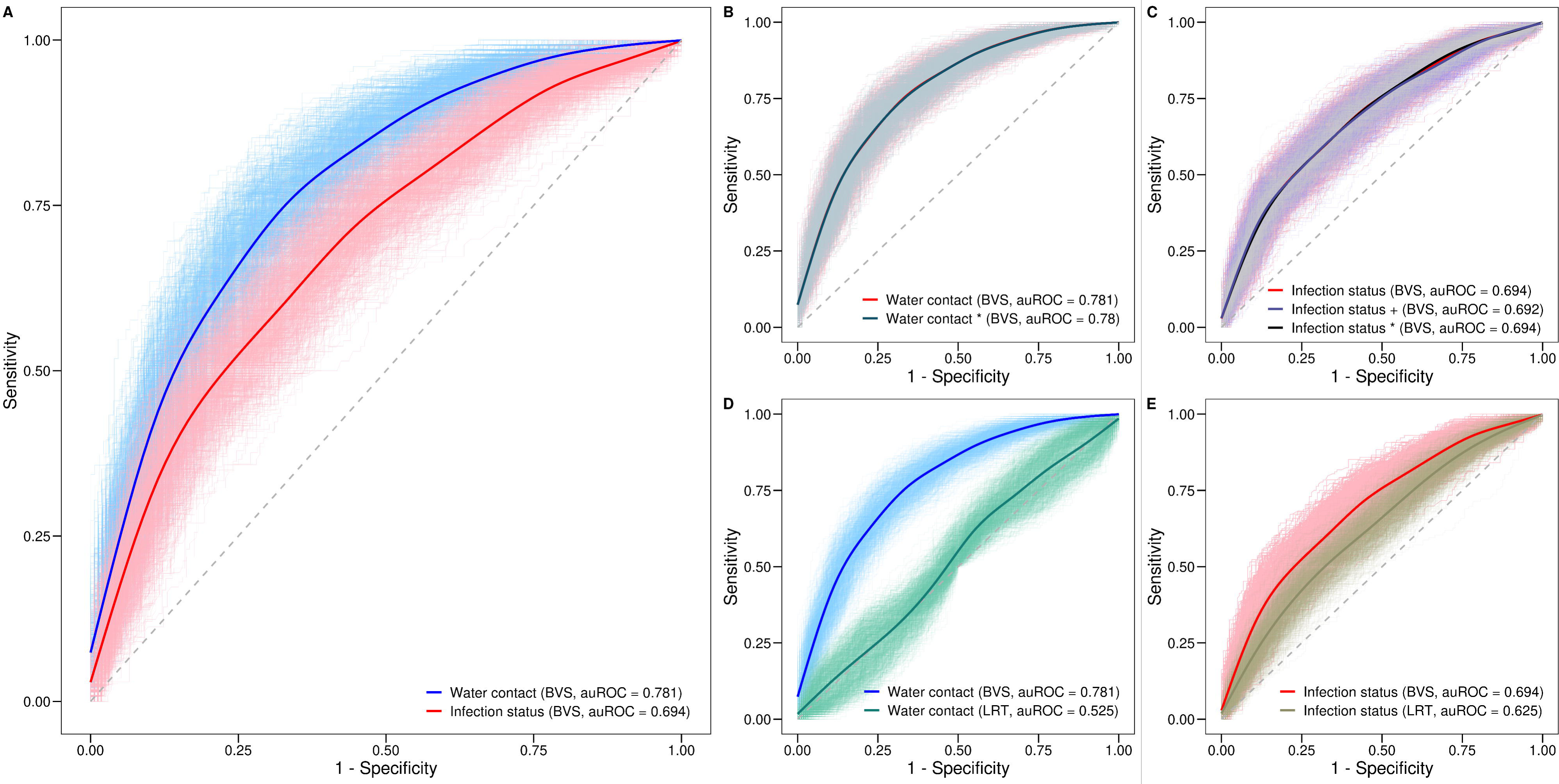
Cross-validated area under the receiver operating curve (auROC) for exposure and infection models. **A.** Comparison of auROC for water contact vs infection status from models in Figs. 7-8 with variables selected using Bayesian variable selection (BVS). **B.** Comparison of water contact model vs water contact model allowing for the selection of additional variables from a set of seven snail variables (water contact * model), implemented using BVS. **C**. Comparison of auROC of infection status model (in Fig. 8) versus auROC of infection status model allowing for the selection of additional variables from a set of 71 granular exposure variables (infection + model) as well as infection model allowing for the selection of additional variables from a set of seven snail variables (infection * model), implemented using BVS. **D.** Comparison of water contact model using BVS versus a water contact model with variables selected using likelihood ratio tests (LRTs). LRTs compared one variable at a time against an ‘empty’ model with village-level fixed effects at p<0.05 to select variables from the same candidate set as BVS. Differences between sets selected via BVS and LRTs are shown in Fig. S11. **E**. Comparison of the infection status model using BVS versus an infection model with variables selected using LRTs. All auROCs were computed using 100 repeats of 10-fold cross-validation. In B-C, additional snail variables selected via BVS were distance to the closest water site with infected snails and the number of infected snails at the closest water site. None of these variables were significant in any model. In C, the six additional granular exposure variables selected were years of residence in the village, time spent on fishing, time spent on washing clothes with soap, household-level fishing, and household-level swimming or playing, and duration of water contact (in hours per week). None of these variables were significant in any model. For D-E, differences between the variable sets selected via BVS and LRTs are shown in Table S8.

## Discussion

WHO guidance for schistosomiasis control has recommended complementing MDA with behaviour change, WASH, or environmental control to reduce transmission [4]. However, as evidence on the effectiveness of complementary interventions is limited, and uncertainties regarding their delivery and integration with MDA persist [25,26], MDA remains the sole strategy applied at scale. Here we conducted a data-driven characterisation of key human-environmental drivers of human schistosome exposure in a sample of 2867 individuals across 38 fishing villages in Uganda. We found that at-risk groups for water contact and infection differed in their observable characteristics suggesting that transmission/exposure and infection control interventions should not be targeted to a single or the same risk group.

Age-specific infection patterns did not approximate age-specific water contact. Despite our population being from three districts with different local climates, geography, tribal and religious groups, water contact was comparatively low in children across all districts. Infection prevalence peaked at age 15—more than a decade earlier than water contact which peaked at age 30. Existing transmission models assume that age-specific water contact is highest in school-age children which relies on a direct correspondence between current age-specific exposure and infection prevalence [22,27]. Yet, we found that correlations between having any current water contact and being currently infected were weak, and that trends in age-dependent infection did not differ significantly based on current water contact. The limited ability of water contact in our study to explain schistosome infection status is consistent with fundamental schistosome biology. Exposure and infection have vastly different timescales. Egg-patent infections can be detected via Kato-Katz microscopy 5-7 weeks after host penetration [28,29] while people can remain infected on average for 7-10 years without exposure [23]. Our findings can inform exposure parameter values for use in future transmission models to reflect empirical trends in exposure. They advocate for relaxed assumptions about exposure-infection correspondence. The role of different exposure measures (past or cumulative exposure) and acquired immunity in explaining infection outcomes needs to be explored in future studies to clarify mechanisms influencing age-specific infection trends.

Gender influenced water contact but was not relevant for predicting infection. After adjustments for covariates such as age, females were more likely to have any water contact when compared to males. When analysed separately by type of activity, females were more likely to engage in domestic water contact activities but less likely to have recreational and occupational water contact than males. However, considering only individuals with any water contact, there were no differences between females and males in the frequency and duration of water contact overall. As gender differences in water contact arose in puberty (ages 15+) and related to occupational status, we found that age and occupation were associated with water contact frequency and duration as well as current infection while gender was not directly related to any of these outcomes. Our findings contrast with a recent meta-analysis by Ayabina et al. [30] reporting gender differences in schistosome infection prevalence, but unlike our study, they were not able to adjust for age or occupation. We suggest that the effects of gender on exposure and infection are directly mediated by independent factors that drive gender differences in exposure but not gender itself. Therefore, future studies should interpret gender effects for infection with caution and avoid extrapolating any such findings to exposure patterns.

Beyond gender, there were substantial differences between risk factors for exposure and infection and, in turn, in the profiles for high-risk groups. Key risk factors for water contact were being an adult (aged 18+), being female, having fishing or fish mongering occupations, and living in villages with more water sites or in villages without safe drinking water. Key risk factors for infection were being a child aged 10-15, having primary-school education, and living in villages with ≥50% infection prevalence. Differences in risk profiles for exposure and infection have implications for screening populations and targeting of interventions. Exposure reduction interventions have been frequently targeted at school-aged children [9,31] which do not necessarily have the highest exposure. Our findings may explain why WASH interventions in randomised controlled trials have failed to reduce infection prevalence or transmission [9]. Recreational water contact has been targeted by past interventions and called an important source of exposure [32,33], but in this study, recreational water contact was relatively unimportant.

WASH interventions for schistosomiasis may need to become more granular as we found that sanitation and water infrastructure differed in their influence on exposure. People living in households close to water sites with visible signs of open defecation had lower water contact which illustrates how individual-level exposure patterns are influenced by collective sanitation behaviour. Availability of sanitation infrastructure did not significantly affect water contact as neither public latrines nor private latrines were selected as predictors of water contact. For safe water, we found that only specific types of water contact were reduced by water infrastructure. In villages where all study households used safe drinking water from taps or boreholes, people had a 69% lower likelihood of having domestic water contact while occupational and recreational water contact were not significantly associated with safe drinking water use. Hence, the benefits of WASH may be activity specific. We also note that while village-level use of safe water was selected, household-level safe water use or the availability of public taps/boreholes were not relevant for predicting exposure. This highlights that both availability of safe water infrastructure and community norms around water use were required for public infrastructure to affect exposure. As females had more domestic water contact than males (75% vs 16%), interventions to reduce domestic water contact may benefit women more than men. In this study, only public not private household-level WASH infrastructure was associated with water contact and WASH was not relevant for predicting current infection as no WASH variables were selected for the infection model. This contrasts with a meta-analysis by Grimes et al. who found significant effects for both household-level and community-level WASH on infection but noted poor study quality, risk of confounding, and inconsistent WASH definitions [12]. Future studies should focus on provision of public infrastructure targeted at specific high-risk behaviours such as washing clothes or fishing and seek to quantify impacts on exposure behaviour.

Hotspots exist in Uganda and elsewhere and are defined as high prevalence areas (≥50% by Kato-Katz microscopy) that are unresponsive to repeated MDA [34,35]. There have been calls for integrated control in these settings, including the use of exposure reduction interventions [36]. Our study was conducted in three districts that each had received over 13 rounds of MDA since the year 2003 [37]. Yet, we found that hotspot villages were associated with lower rates of water contact when compared to villages with <50% infection prevalence. This suggests that hotspots were not characterised by more intense exposure patterns.

Distance to waterbodies has been widely used to predict infection risk, approximate the force of infection, determine sampling in infection studies, and decide where to implement MDA [14,20,38,39]. Past studies have used binary indicators of distance with varying cut-offs ranging from 0.5-5 km to predict infection [20,21,39–41]. The rationale for these cut-offs and the mechanism through which distance influences infection likelihood are unclear. We found strong exposure in household-level exposure gradients within small geographical areas; the prevalence of water contact declined by an absolute 3.4% for every 100m-increase in household distance to the closest water site between 0-1 km. By contrast, infection prevalence showed a weaker gradient declining only by 1.9% between 0-1 km household distance. Studies have used different distance measures such as household distance [21,40,41], or village distance [42]. We showed that these measures were not interchangeable as we found no exposure gradients in school-based measures. We also found that school enrolment was not relevant for predicting water contact. These findings suggest that school-based interventions, although appropriate for MDA and targeting high-risk infection groups, may be ineffective for exposure which is further emphasised by lower rates of water contact in children versus adults.

The existence of exposure gradients over waterbody distance to households also raises the possibility of different reinfection trends over distance. If such fine-scale reinfection trends exist, household or a cruder approximation of village distance based on water contact patterns should be incorporated into algorithms for selecting the number of rounds of MDA. The optimal spatial units for targeting MDA are still debated [20] because the empirical size of transmission units remains to be established. Collecting and analysing fine-scale exposure data could play an important role for identifying spatial units of transmission and devising MDA strategies targeted at these units.

A key challenge for studies has been reliable exposure measurement. Self-reported measures have been the most widely used [3] but are considered less reliable than direct observations or wearable global positioning system (GPS) logger data [43,44]. Here, we triangulated self-reported water contact with direct observations, demonstrating that surveys provide a cost-effective way to obtain valid population-level exposure measures. We note however, that while we found a good representation of overall community-level trends in self-reported data, some behaviours such as recreational water contact of children could be underreported as water contact was reported by the household head. It remains possible that children are sent to retrieve drinking water, hence reporting this activity, but also go for a leisurely swim which was not reported. To address some aspects of exposure that are difficult to capture using self-reports, research is currently underway in SchistoTrack to produce more granular measurement tools based on wearable GPS loggers. With finer spatial measurements, it may be possible to better capture more exact durations of water contact, intensity of exposure, seasonal and diurnal exposure patterns, and explore the role of human mobility for schistosome transmission.

We conducted a comprehensive characterisation of schistosome exposure and found that determinants of current water contact were different from determinants of current infection. Our findings on exposure risk profiles enable better targeting and gender equity of WASH interventions to support integrated control of schistosomiasis. With further precision mapping of exposure, MDA treatment may be optimised based on waterbody distance to reach groups with the highest exposure while avoiding over-treatment of lower risk groups further away from open waterbodies. Past behaviour change interventions have frequently targeted children, but future research is needed for interventions targeted at adults and using community-level WASH to reduce schistosome exposure.

## Methods

### Study setting and sampling

This was a cross-sectional study in Uganda using baseline data from the SchistoTrack Study cohort [24], a collaboration between the University of Oxford and the Uganda Ministry of Health. Data was collected in January and February 2022 in 38 rural fishing villages across three districts (Pakwach, Buliisa, and Mayuge). We sampled villages within 3 km distance (based on the village centre location reported by the local chairman) of the shorelines of either the River Nile, Lake Albert or Lake Victoria (Fig. 2b-d). In all study districts, MDA using praziquantel has been carried out since 2003, targeting all children aged 5+. Thirteen or more rounds of treatment have been administered to-date, the most recent having been conducted over one year before this study. We randomly sampled a total of 1459 households, approximately 40 per village, from village registers or MDA records. All households with at least one child and one adult residing in the village for at least six months of the year were eligible. After obtaining informed consent, questionnaires were administered to obtain socio-demographics, biomedical variables, WASH and environmental variables, and water contact patterns on all household members aged 1+. At the end of the interview, one adult (aged 18+) and one child (aged 5-17) per household were selected for clinical assessments by the household head. Fig. 1 depicts the participant flow resulting in an analytical sample of 2867 participants from 1444 households. A more detailed participant flow diagram is shown in Fig. S12. This study was designed to detect a minimum effect size of 8% with an unevenly exposed population and a household design effect of 1.136 at 97.5% power.

We also conducted village-level infrastructure and malacology surveys. For village-level infrastructure, we collected information on the number of public latrines and public taps/boreholes and their respective GPS locations, and the number of roads per village as well as the GPS locations of all primary schools. To assess environmental risk, malacologists mapped all water sites in the study area, guided by the village chairman or a village health worker, and recorded GPS points, site type, and the observed presence of human faeces of all water sites within the village. Malacologists collected all living and recently dead snails at each water site for 30 minutes. Snail infectivity was established by exposing snails to natural sunlight and determining the number of snails shedding human schistosome cercaria by water site.

### S. mansoni infection

Kato-Katz stool microscopy and point-of-care cathodic antigen (POC-CCA) diagnoses were completed for schistosomiasis as described in Anjorin et al. [37]. Readings were converted to eggs per gram (EPG) of stool. Infection was defined as >0 EPG. In accordance with the current WHO treatment guideline [4], heavy infection intensity was defined as ≥400 EPG. We chose infection status (based on Kato-Katz diagnosis) as the primary infection measure as infection status is relevant for transmission and prevalence has been used to monitor reductions in transmission [45] and identify persistent hotspots [46,47]. POC-CCA was not chosen as the primary infection measure due to the lack of specificity [48] and undemonstrated relevance for transmission [49]. POC-CCA only was used in sensitivity analyses to demonstrate the robustness of main models in the potential scenario of missed light infection intensities.

### Exposure

Survey questions were administered to the household head and/or wife to elicit information on 11 different water contact activities conducted by household members, and their typical weekly frequency, duration, and time of day. The choice of activities was informed by local collaborators to ensure all common water contact activities in the study area were captured. Recorded activities were washing clothes with soap, washing clothes without soap, getting drinking water, washing jerry cans or household items, fishing, fish mongering, swimming or playing, bathing with soap, bathing without soap, collecting papyrus, and collecting shells. For each activity the typical number of trips per household per week was elicited. The typical duration per trip to the water for each activity in the household was elicited using pre-defined duration categories (less than 30 minutes, 1 hour, 2 hours, 3 hours, 4 hours or more). The typical time of day for each activity was elicited using the following categories: before sunrise (1-6am), after sunrise (6-9 am), late morning (9 am-12 pm), midday (12-3 pm), late afternoon (3-5 pm), early evening (5-7 pm), after sunset (7-9 pm), late evening (9 pm-1 am).

Our main exposure outcome was having water contact and it was coded as binary and defined as an individual engaging at least in one water contact activity. We chose having water contact as the primary exposure outcome based on a recent meta-analysis which showed that having any water contact was associated with infection status [3]. Secondary outcomes included the type of water contact activity and frequency and duration of water contact. For activity types, we grouped water contact into three distinct categories: domestic, occupational, and recreational water contact. Domestic water contact was defined as engaging in getting drinking water, washing clothes with/without soap, bathing with/without soap, washing jerry cans or household items, or washing clothes. Occupational water contact was defined as engaging in collecting papyrus, fishing, fish mongering or collecting shells. Recreational water contact was defined as swimming or playing. To obtain individual-level water contact frequency, we summed the self-reported number of water contacts across all activities for each person. Duration was obtained by multiplying the frequency of each activity with its duration and summing durations across all activities for each person.

### Covariates

Detailed definitions of all variables are provided in Table S1. We curated a set of 36 candidate variables for our main analysis and aimed at comprehensively covering key human-environmental determinants of exposure and infection (30 variables for exposure models and six additional exposure variables for the infection models only). At the individual level, we included age, gender, occupation, and education as covariates, based on their relevance for exposure and infection [13,21,50,51]. Age was coded to the nearest year and was a discrete variable as grouping age into categories could lead to information loss [52]. We generated age^2^, age^3^, and age^4^ terms to model nonlinear associations of exposure and infection with age using polynomials. To account for the influence of socio-economic status, we generated a home quality score variable where the quality of the roof, wall, and floor for each household were ranked from 1-4 and summed. Following Chami et al., the rank order of the materials in ascending order was grass, sticks, plastic, and metal for the roof; mud and sticks, plastic, metal, and bricks or cement for the walls; mud, plastic, wood planks, and brick or cement for the floor [5]. WASH access may influence exposure via crowding out open water contact or may affect infection via reducing contamination and thus environmental risk [3,12]. We generated WASH variables indicating the presence of a home latrine and safe drinking water which corresponded to improved sanitation and improved drinking water, respectively, as per WHO-UNICEF joint monitoring committee (JMP) 2017 methodology guideline definitions [53]. Having a flush or pour flush toilet, a covered pit latrine with/without privacy, or a composting toilet were considered as having a home latrine. Households who reported open defecation or having only bucket latrines were considered as not having a safe private home latrine. For drinking water, we considered use of piped water, village taps, boreholes, protected/unprotected dugs, wells, or springs, or rainwater tanks as safe, as per WHO-UNICEF JMP guidelines [53]. Water taken directly from a lake, river or swamp was considered as an unsafe source. We also calculated the village-level prevalence of safe drinking water use and generated variables for the number of latrines, and number of taps/boreholes per village. The number of roads and the type of water site in each village were collected by the study team using a village-level survey. Number of water sites per village were calculated from water sites mapped by malacologists. Past research has shown the existence of lake-gradients in infection [21,41]. We therefore generated Euclidean distance variables to assess whether these gradients were explained by variation in exposure behaviour across lake distance. Variables were defined as follows. Household lake distance was the distance of the household GPS location to the closest water site mapped by malacologists. Village lake distance was computed based on assigning households to the village centre reported by the local chairman and calculating the Euclidean distance to the closest water site. For school lake distance, all children attending primary school were assigned the school closest to the household. We then calculated the Euclidean distance from the school to the closest water site. Distance to the type of water site closest to the household was generated using information on the type of site (river, marsh, beach, swamp, pond) collected by malacologists. Village-level prevalence was defined as per WHO guidelines on the control and elimination of schistosomiasis with 10–49% prevalence by Kato-Katz corresponding to moderate and ≥50% prevalence corresponding to high endemicity [4]. There were no low-prevalence (<10%) villages in our study. For models shown in Fig. 9b-c, we included additional snail variables and water contact variables. The snail variables included the number of water sites with infected snails per village, number of snails within 1 km of the household, number of snails per village, number of infected snails per village, presence of any infected snails in the village, distance to closest water site with infected snails, and whether the closest infected water site from the household was within the village. For water contact, 82 granular water contact variables indicating the frequency and duration for each of the 11 water contact activities, the time of day and whether an activity was conducted at a high-risk time (10 am-3pm) for each activity, were included. We also included variables for the frequency and duration of domestic, recreational, and domestic activities and variables indicating whether anyone in the household conducted any of the 11 water contact activities.

### Comparison of self-reported water contact with direct water contact observations

Direct water contact observations were conducted in a subset of 12 villages concurrently with the collection of self-reported exposure data. All water contacts at two water sites per village were recorded by trained local community village health team members who were trusted by the communities and knew all individuals within their village. Contact was observed between 6 am-6 pm for two weeks and each visitation of the water site, defined having at least one body part immersed, was recorded as a separate water contact event. Age group, as defined per recent WHO guidelines on the control and elimination of schistosomiasis [4] as well as gender, activity, bodily immersion, time, and duration were recorded. By comparing the age distribution in direct observation data for participants aged 5+ with the age distribution of water contact in self-reported data from the same 12 villages, we assessed the extent to which survey data was representative of the age composition in the observation data.

### Geospatial data

Waterbody boundaries shown in Fig. 2 are from a public shapefile of waterbodies in Uganda, made available online under a Creative Commons Attribution (Non-commercial 4.0 International) by the World Resources Institute [54].

### Statistical analysis

All statistical analyses were conducted in R version 4.1.0. To model non-linear trends of exposure and infection over age, we used generalised additive mixed models (GAMs) from the mgcv package [55]. The number of knots were separately chosen for each GAM based on the mgcv diagnostic function ‘k.check’. We obtained distance gradients from GAMs by computing the average slope over household distance between 0-1 km. We refrained from reporting distance gradients >1 km household distance, as only 4.7% of participants (137/2867) lived in household >1 km from a water site. We used Pearson correlation coefficients to assess correlations between water contact and infection status across participants. We also assessed pairwise correlations of these variables within households. All t-tests conducted in this paper were two-sided. To select predictors of water contact and infection status, respectively, the same variable selection procedure was applied separately using the same list of input covariates (except for six additional exposure variables which were only included in the infection status models). To assess the relevance of snail variables for exposure and infection as well as the relevance of more granular exposure variables for predicting infection, we used Bayesian variable selection to select additional predictors from the sets of seven snail variables and 82 granular exposure variables, respectively, while constraining all previously selected variables to remain included. This way, we tested whether adding snail variables or more granular exposure variables improved predictions compared to the models with our main sets of predictor variables. We implemented Bayesian variable selection using the BAS package in R [56]. The outputs of this analysis were a ranking of the best-fitting models (with associated posterior probabilities for each model being the best model) as well as marginal inclusion probabilities (bounded between 0 and 1) for each variable, indicating the importance of including each of the predictors for exposure and infection models, respectively. The large number of candidate variables yielded a prohibitively large model space (2^30^ possible models). Therefore, we used Markov Chain Monte Carlo (MCMC) methods implemented within the BAS package to sample models based on their posterior probabilities which avoided enumerating all possible models [57]. We ran 2*10^8^ MCMC iterations, a number deemed sufficient based on in-built MCMC diagnostics of the BAS package. The prior of the model distribution was set to a uniform distribution which assigns equal probabilities to all models. For the distributions on the regression coefficients, we used Jeffreys-Zellner-Siow-priors which have been shown to have desirable mathematical properties and provide more consistent results under both the null and the alternative model than alternative priors such as hyper-g priors or Empirical Bayes priors and are also more consistent than non-Bayesian methods such as Akaike information criterion [58–60]. To build the final exposure and infection models, we selected all variables with inclusion probabilities of pr ≥0.5 in the Bayesian variable selection output which yields the so-called median probability model [61]. The choice of pr ≥0.5 was motivated by research showing that the median probability model, not the highest probability model, often yields the best predictive performance [57]. As an alternative to Bayesian variable selection, we used LRTs with a cut-off of p < 0.05. For LRTs we compared a model with just one variable at the time against an empty model with village-level fixed effects only to select predictors of exposure and infection. A comparison of the variable sets selected using Bayesian variable selection and LRTs is shown in Table S8.

After variable selection, we used logistic regression models to predict whether an individual had any water contact (n=2867). For all regression models, we clustered standard errors at the household level to account for our sampling design where households were randomly selected, but not individuals. To identify which variables predicted a specific type of water contact, we used the main set of exposure variables to predict domestic, occupational, and recreational water contact using separate logistic regression models. In these models, we removed all participants engaging in multiple types of water contact activities to generate mutually exclusive categories. This removal resulted in 6.3% individuals (183/2867) being excluded. To adequately model overdispersion of water contact frequency and duration, we used negative binomial models to predict frequency and duration among participants with water contact (1339/2867) using all selected predictors of exposure. For infection, we used the selected infection variables in logistic regression models to predict infection status as well as heavy infection (n=2867). Across all models, we computed generalised variance inflation factors (gVIFs) to diagnose multicollinearity but did not remove any variables as none had gVIFs >10. Ten-fold cross-validation was used to assess predictive power of the statistical models.

## Data availability

Data is not publicly available due to data protection and ethics restrictions related to the ongoing nature of the SchistoTrack Cohort and easily identifiable nature of the participants.

## Ethics approval

Data collection and use were reviewed and approved by Oxford Tropical Research Ethics Committee (OxTREC 509-21), Vector Control Division Research Ethics Committee of the Uganda Ministry of Health (VCDREC146), and Uganda National Council of Science and Technology (UNCST HS 1664ES).

## Acknowledgements

We are thankful for the involvement of our study participants and local communities. We thank all field teams including malacologists, surveyors, nurses, and laboratory technicians involved in the 2022 SchistoTrack baseline data collection, especially Benjamin Tinkitina who led the household survey team. The support of the Uganda Ministry of Health, local district leaders, focal health workers, and village health teams was crucial for building partnerships and continued trust with study communities.

## Author contributions

Conceptualization: F.R. and G.F.C. Data curation: F.R., N.B.K., A.M.B., F.B., B.N., and G.F.C. Formal analysis: F.R. Funding acquisition: G.F.C. Investigation: F.R. and G.F.C. Methodology: F.R. and G.F.C. Project administration: N.B.K. and G.F.C. Resources: N.B.K. and G.F.C. Software: G.F.C. Supervision: G.F.C. Validation: F.R. Visualization: F.R. Writing – original draft: F.R. Writing – review & editing: F.R., N.B.K., A.M.B., F.B., B.N., and G.F.C.

## Conflicts of interest

The authors declare no conflicts of interest.

## Funding

A DPhil scholarship was awarded from the Nuffield Department of Population Health (NDPH) to Fabian Reitzug. Grants from the Wellcome Trust Institutional Strategic Support Fund (204826/Z/16/Z), NDPH Pump Priming Fund, John Fell Fund, Robertson Foundation Fellowship, and UKRI EPSRC Award (EP/X021793/1) were awarded to Goylette F. Chami.

## Open access

For the purpose of Open Access, the author has applied a CC-BY public copyright licence to any Author Accepted Manuscript version arising from this submission.

## 1 Supplementary Tables

**Table S1:**
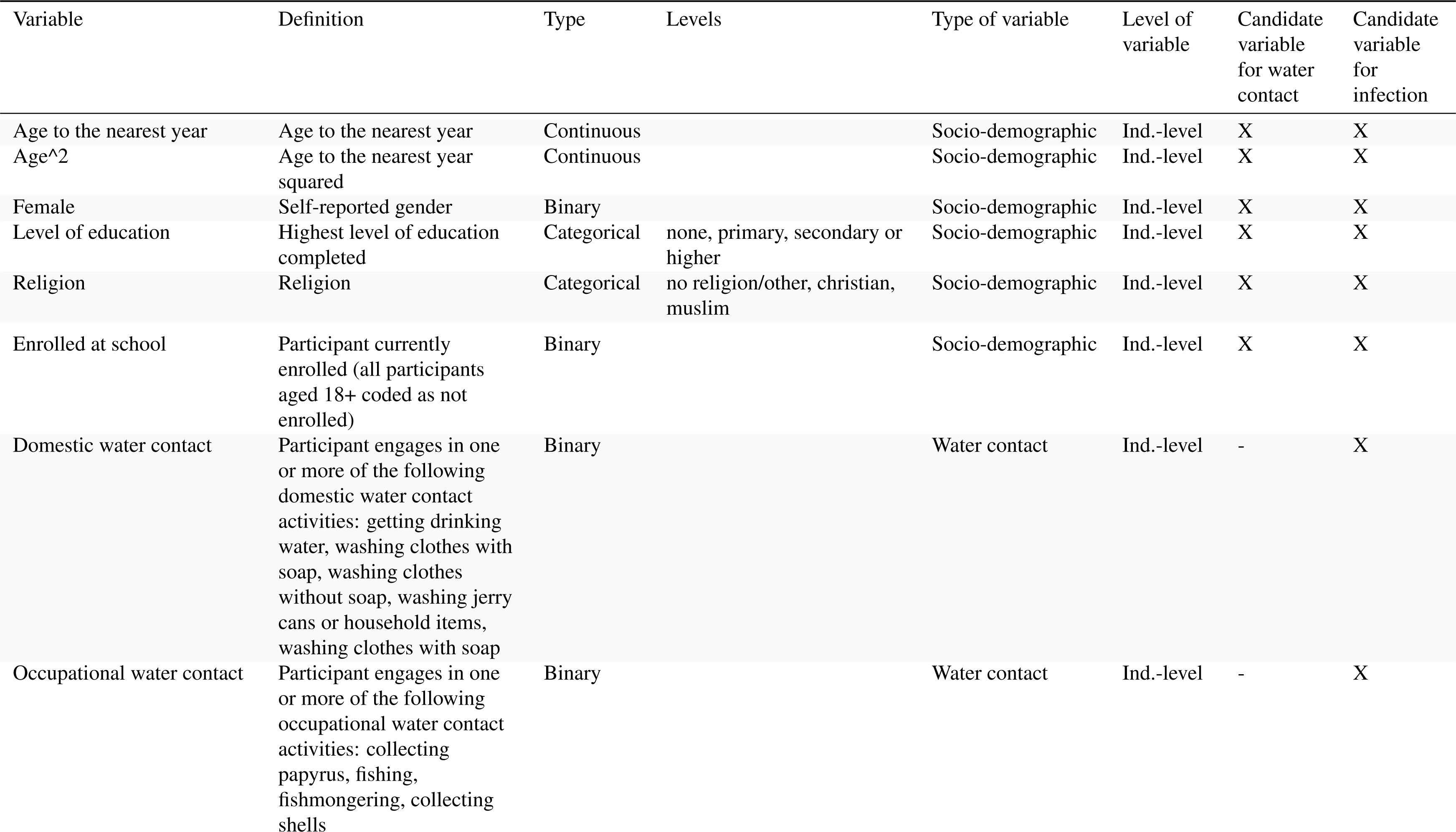
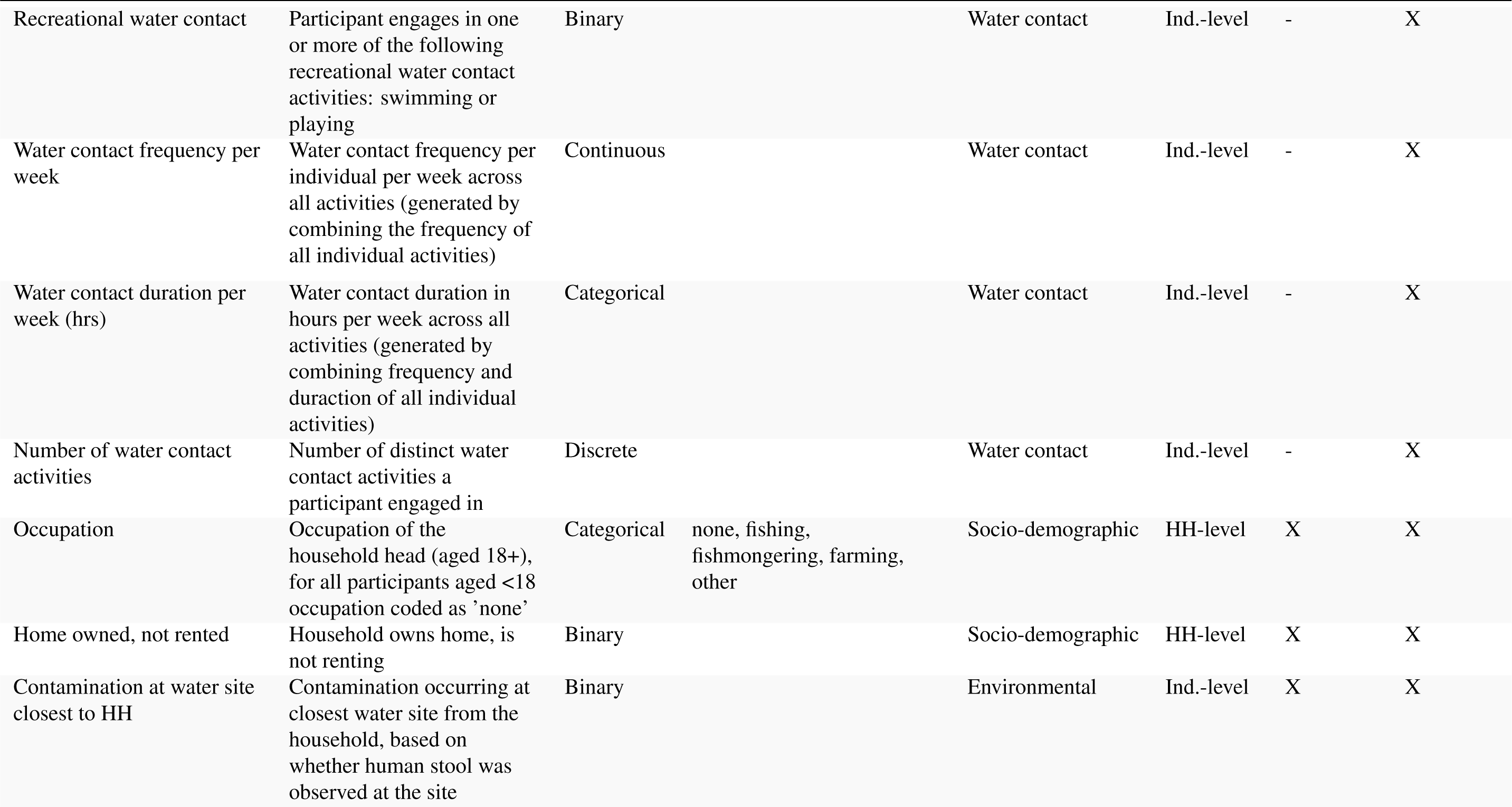
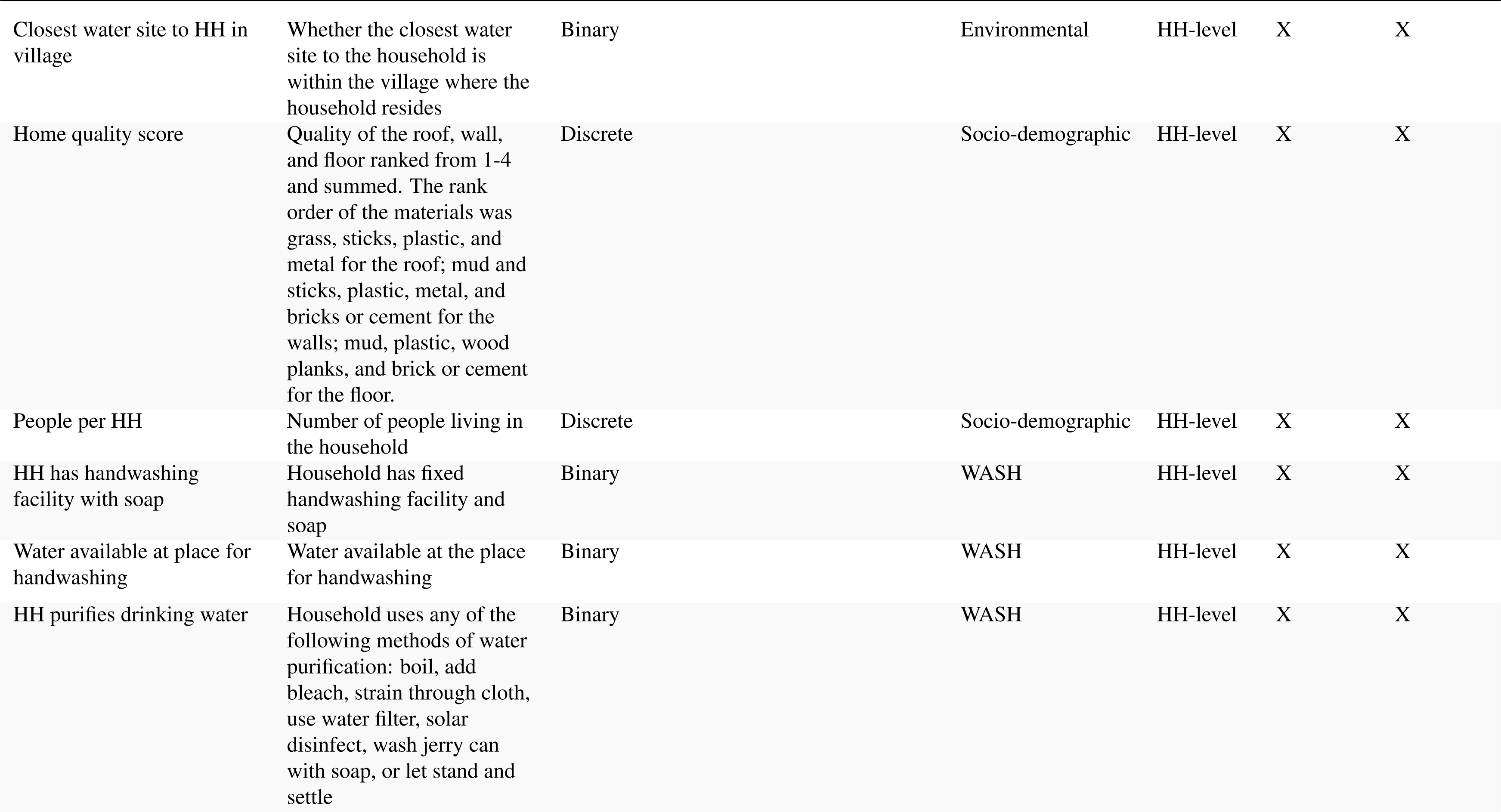
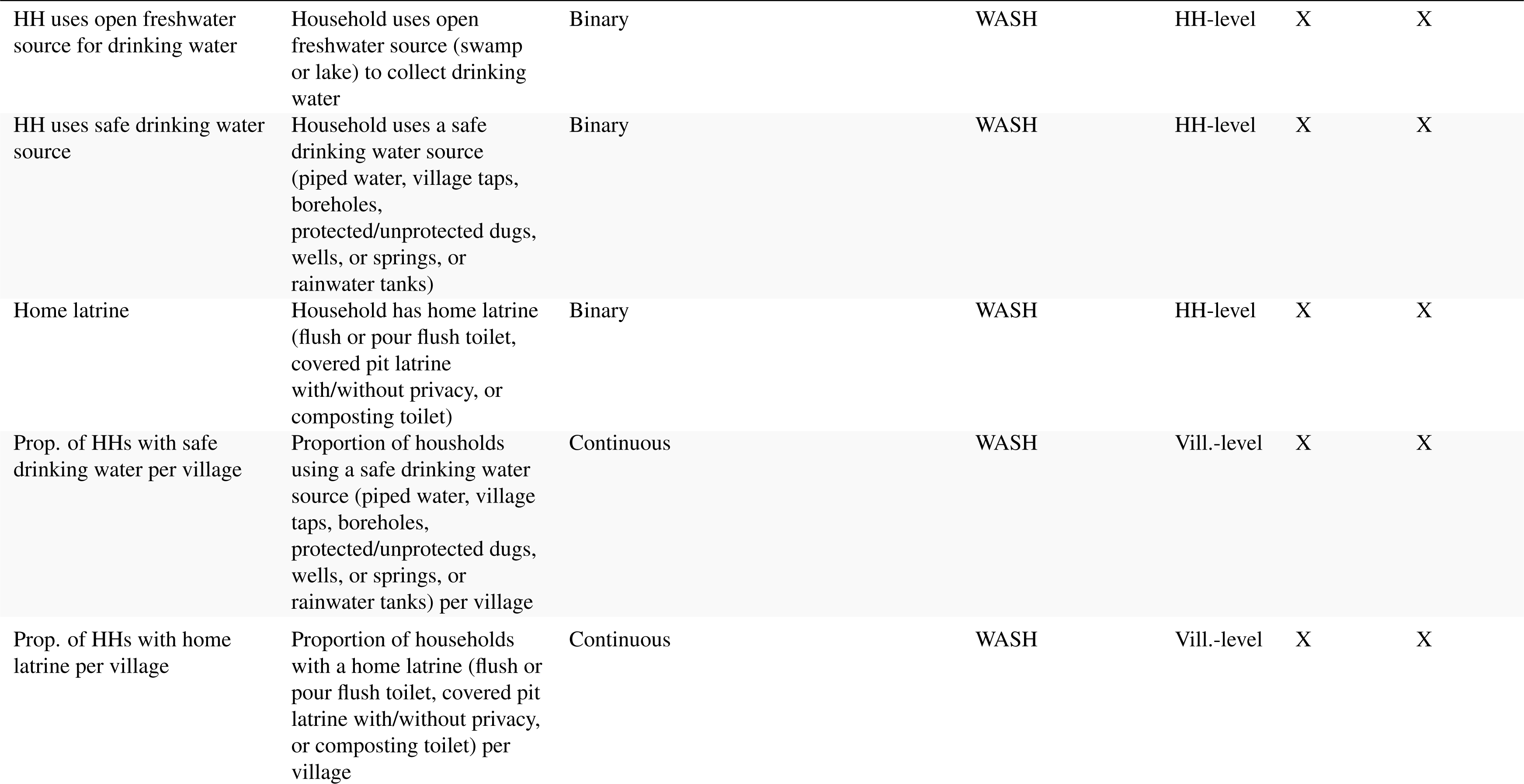
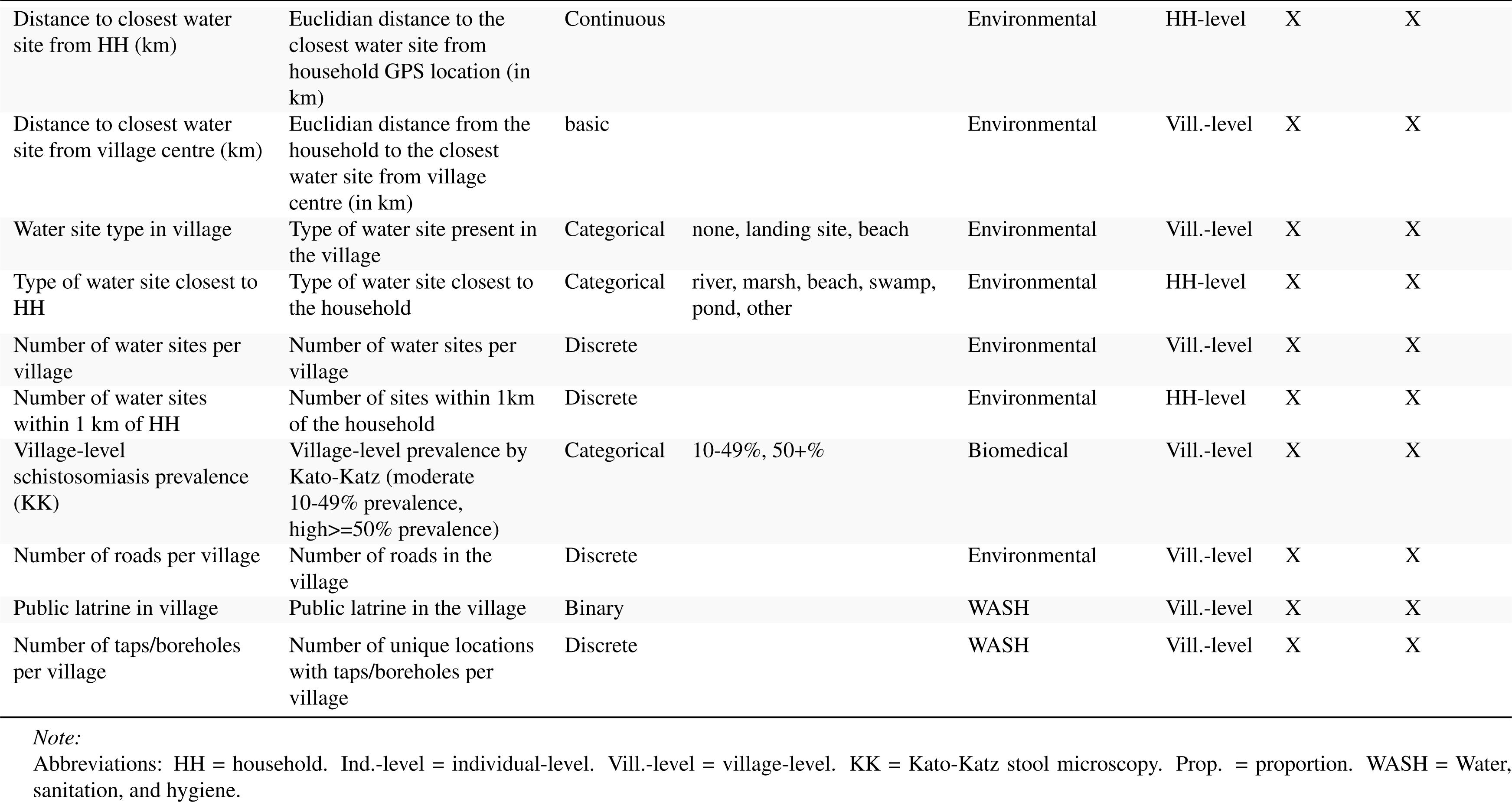
Variable definitions.

| Variable | Definition | Type | Levels | Type of variable | Level of variable | Candidate variable for water contact | Candidate variable for infection |
| --- | --- | --- | --- | --- | --- | --- | --- |
| Age to the nearest year | Age to the nearest year | Continuous |  | Socio-demographic | Ind.-level | X | X |
| Age^2 | Age to the nearest year squared | Continuous |  | Socio-demographic | Ind.-level | X | X |
| Female | Self-reported gender | Binary |  | Socio-demographic | Ind.-level | X | X |
| Level of education | Highest level of education completed | Categorical | none, primary, secondary or higher | Socio-demographic | Ind.-level | X | X |
| Religion | Religion | Categorical | no religion/other, christian, muslim | Socio-demographic | Ind.-level | X | X |
| Enrolled at school | Participant currently enrolled (all participants aged 18+ coded as not enrolled) | Binary |  | Socio-demographic | Ind.-level | X | X |
| Domestic water contact | Participant engages in one or more of the following domestic water contact activities: getting drinking water, washing clothes with soap, washing clothes without soap, washing jerry cans or household items, washing clothes with soap | Binary |  | Water contact | Ind.-level | - | X |
| Occupational water contact | Participant engages in one or more of the following occupational water contact activities: collecting papyrus, fishing, fishmongering, collecting shells | Binary |  | Water contact | Ind.-level | - | X |
| Recreational water contact | Participant engages in one or more of the following recreational water contact activities: swimming or playing | Binary |  | Water contact | Ind.-level | - | X |
| Water contact frequency per week | Water contact frequency per individual per week across all activities (generated by combining the frequency of all individual activities) | Continuous |  | Water contact | Ind.-level | - | X |
| Water contact duration per week (hrs) | Water contact duration in hours per week across all activities (generated by combining frequency and duration of all individual activities) | Categorical |  | Water contact | Ind.-level | - | X |
| Number of water contact activities | Number of distinct water contact activities a participant engaged in | Discrete |  | Water contact | Ind.-level | - | X |
| Occupation | Occupation of the household head (aged 18+), for all participants aged <18 occupation coded as 'none' | Categorical | none, fishing, fishmongering, farming, other | Socio-demographic | HH-level | X | X |
| Home owned, not rented | Household owns home, is not renting | Binary |  | Socio-demographic | HH-level | X | X |
| Contamination at water site closest to HH | Contamination occurring at closest water site from the household, based on whether human stool was observed at the site | Binary |  | Environmental | Ind.-level | X | X |
| Closest water site to HH in village | Whether the closest water site to the household is within the village where the household resides | Binary |  | Environmental | HH-level | X | X |
| Home quality score | Quality of the roof, wall, and floor ranked from 1-4 and summed. The rank order of the materials was grass, sticks, plastic, and metal for the roof; mud and sticks, plastic, metal, and bricks or cement for the walls; mud, plastic, wood planks, and brick or cement for the floor. | Discrete |  | Socio-demographic | HH-level | X | X |
| People per HH | Number of people living in the household | Discrete |  | Socio-demographic | HH-level | X | X |
| HH has handwashing facility with soap | Household has fixed handwashing facility and soap | Binary |  | WASH | HH-level | X | X |
| Water available at place for handwashing | Water available at the place for handwashing | Binary |  | WASH | HH-level | X | X |
| HH purifies drinking water | Household uses any of the following methods of water purification: boil, add bleach, strain through cloth, use water filter, solar disinfect, wash jerry can with soap, or let stand and settle | Binary |  | WASH | HH-level | X | X |
| HH uses open freshwater source for drinking water | Household uses open freshwater source (swamp or lake) to collect drinking water | Binary |  | WASH | HH-level | X | X |
| HH uses safe drinking water source | Household uses a safe drinking water source (piped water, village taps, boreholes, protected/unprotected dug wells, or springs, or rainwater tanks) | Binary |  | WASH | HH-level | X | X |
| Home latrine | Household has home latrine (flush or pour flush toilet, covered pit latrine with/without privacy, or composting toilet) | Binary |  | WASH | HH-level | X | X |
| Prop. of HHs with safe drinking water per village | Proportion of households using a safe drinking water source (piped water, village taps, boreholes, protected/unprotected dug wells, or springs, or rainwater tanks) per village | Continuous |  | WASH | Vill.-level | X | X |
| Prop. of HHs with home latrine per village | Proportion of households with a home latrine (flush or pour flush toilet, covered pit latrine with/without privacy, or composting toilet) per village | Continuous |  | WASH | Vill.-level | X | X |
| Distance to closest water site from HH (km) | Euclidian distance to the closest water site from household GPS location (in km) | Continuous |  | Environmental | HH-level | X | X |
| Distance to closest water site from village centre (km) | Euclidian distance from the household to the closest water site from village centre (in km) | basic |  | Environmental | Vill.-level | X | X |
| Water site type in village | Type of water site present in the village | Categorical | none, landing site, beach | Environmental | Vill.-level | X | X |
| Type of water site closest to HH | Type of water site closest to the household | Categorical | river, marsh, beach, swamp, pond, other | Environmental | HH-level | X | X |
| Number of water sites per village | Number of water sites per village | Discrete |  | Environmental | Vill.-level | X | X |
| Number of water sites within 1 km of HH | Number of sites within 1km of the household | Discrete |  | Environmental | HH-level | X | X |
| Village-level schistosomiasis prevalence (KK) | Village-level prevalence by Kato-Katz (moderate 10-49% prevalence, high>=50% prevalence) | Categorical | 10-49%, 50+% | Biomedical | Vill.-level | X | X |
| Number of roads per village | Number of roads in the village | Discrete |  | Environmental | Vill.-level | X | X |
| Public latrine in village | Public latrine in the village | Binary |  | WASH | Vill.-level | X | X |
| Number of taps/boreholes per village | Number of unique locations with taps/boreholes per village | Discrete |  | WASH | Vill.-level | X | X |

*Note:*
Abbreviations: HH = household. Ind.-level = individual-level. Vill.-level = village-level. KK = Kato-Katz stool microscopy. Prop. = proportion. WASH = Water, sanitation, and hygiene.

**Table S2:**
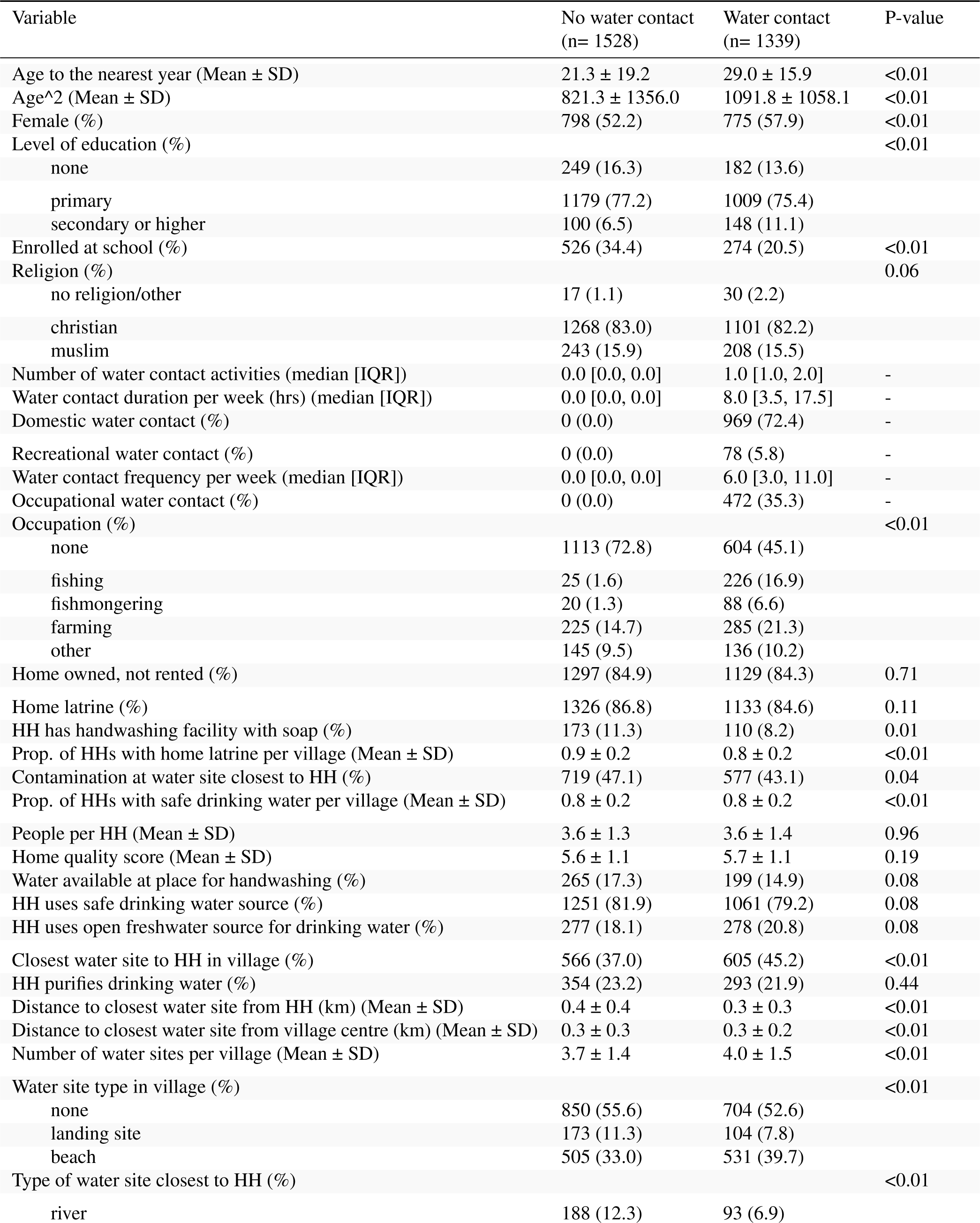
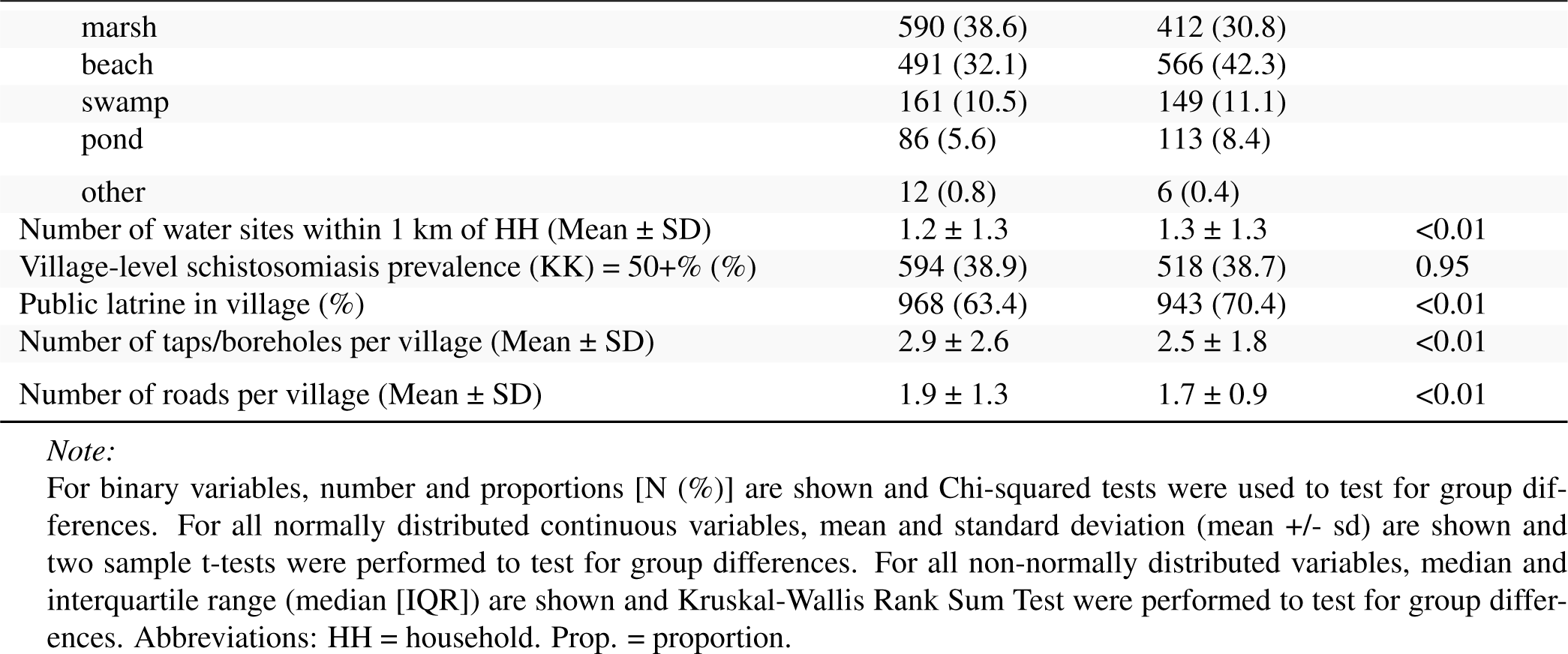
Key participant characteristics.

| Variable | No water contact<br>(n= 1528) | Water contact<br>(n= 1339) | P-value |
| --- | --- | --- | --- |
| Age to the nearest year (Mean ± SD) | 21.3 ± 19.2 | 29.0 ± 15.9 | <0.01 |
| Age<sup>2</sup> (Mean ± SD) | 821.3 ± 1356.0 | 1091.8 ± 1058.1 | <0.01 |
| Female (%) | 798 (52.2) | 775 (57.9) | <0.01 |
| Level of education (%) |  |  | <0.01 |
| none | 249 (16.3) | 182 (13.6) |  |
| primary | 1179 (77.2) | 1009 (75.4) |  |
| secondary or higher | 100 (6.5) | 148 (11.1) |  |
| Enrolled at school (%) | 526 (34.4) | 274 (20.5) | <0.01 |
| Religion (%) |  |  | 0.06 |
| no religion/other | 17 (1.1) | 30 (2.2) |  |
| christian | 1268 (83.0) | 1101 (82.2) |  |
| muslim | 243 (15.9) | 208 (15.5) |  |
| Number of water contact activities (median [IQR]) | 0.0 [0.0, 0.0] | 1.0 [1.0, 2.0] | - |
| Water contact duration per week (hrs) (median [IQR]) | 0.0 [0.0, 0.0] | 8.0 [3.5, 17.5] | - |
| Domestic water contact (%) | 0 (0.0) | 969 (72.4) | - |
| Recreational water contact (%) | 0 (0.0) | 78 (5.8) | - |
| Water contact frequency per week (median [IQR]) | 0.0 [0.0, 0.0] | 6.0 [3.0, 11.0] | - |
| Occupational water contact (%) | 0 (0.0) | 472 (35.3) | - |
| Occupation (%) |  |  | <0.01 |
| none | 1113 (72.8) | 604 (45.1) |  |
| fishing | 25 (1.6) | 226 (16.9) |  |
| fishmongering | 20 (1.3) | 88 (6.6) |  |
| farming | 225 (14.7) | 285 (21.3) |  |
| other | 145 (9.5) | 136 (10.2) |  |
| Home owned, not rented (%) | 1297 (84.9) | 1129 (84.3) | 0.71 |
| Home latrine (%) | 1326 (86.8) | 1133 (84.6) | 0.11 |
| HH has handwashing facility with soap (%) | 173 (11.3) | 110 (8.2) | 0.01 |
| Prop. of HHs with home latrine per village (Mean ± SD) | 0.9 ± 0.2 | 0.8 ± 0.2 | <0.01 |
| Contamination at water site closest to HH (%) | 719 (47.1) | 577 (43.1) | 0.04 |
| Prop. of HHs with safe drinking water per village (Mean ± SD) | 0.8 ± 0.2 | 0.8 ± 0.2 | <0.01 |
| People per HH (Mean ± SD) | 3.6 ± 1.3 | 3.6 ± 1.4 | 0.96 |
| Home quality score (Mean ± SD) | 5.6 ± 1.1 | 5.7 ± 1.1 | 0.19 |
| Water available at place for handwashing (%) | 265 (17.3) | 199 (14.9) | 0.08 |
| HH uses safe drinking water source (%) | 1251 (81.9) | 1061 (79.2) | 0.08 |
| HH uses open freshwater source for drinking water (%) | 277 (18.1) | 278 (20.8) | 0.08 |
| Closest water site to HH in village (%) | 566 (37.0) | 605 (45.2) | <0.01 |
| HH purifies drinking water (%) | 354 (23.2) | 293 (21.9) | 0.44 |
| Distance to closest water site from HH (km) (Mean ± SD) | 0.4 ± 0.4 | 0.3 ± 0.3 | <0.01 |
| Distance to closest water site from village centre (km) (Mean ± SD) | 0.3 ± 0.3 | 0.3 ± 0.2 | <0.01 |
| Number of water sites per village (Mean ± SD) | 3.7 ± 1.4 | 4.0 ± 1.5 | <0.01 |
| Water site type in village (%) |  |  | <0.01 |
| none | 850 (55.6) | 704 (52.6) |  |
| landing site | 173 (11.3) | 104 (7.8) |  |
| beach | 505 (33.0) | 531 (39.7) |  |
| Type of water site closest to HH (%) |  |  | <0.01 |
| river | 188 (12.3) | 93 (6.9) |  |
| marsh | 590 (38.6) | 412 (30.8) |  |
| beach | 491 (32.1) | 566 (42.3) |  |
| swamp | 161 (10.5) | 149 (11.1) |  |
| pond | 86 (5.6) | 113 (8.4) |  |
| other | 12 (0.8) | 6 (0.4) |  |
| Number of water sites within 1 km of HH (Mean ± SD) | 1.2 ± 1.3 | 1.3 ± 1.3 | <0.01 |
| Village-level schistosomiasis prevalence (KK) = 50+ % (%) | 594 (38.9) | 518 (38.7) | 0.95 |
| Public latrine in village (%) | 968 (63.4) | 943 (70.4) | <0.01 |
| Number of taps/boreholes per village (Mean ± SD) | 2.9 ± 2.6 | 2.5 ± 1.8 | <0.01 |
| Number of roads per village (Mean ± SD) | 1.9 ± 1.3 | 1.7 ± 0.9 | <0.01 |

*Note:*
For binary variables, number and proportions [N (%)] are shown and Chi-squared tests were used to test for group differences. For all normally distributed continuous variables, mean and standard deviation (mean ± sd) are shown and two sample t-tests were performed to test for group differences. For all non-normally distributed variables, median and interquartile range (median [IQR]) are shown and Kruskal-Wallis Rank Sum Test were performed to test for group differences. Abbreviations: HH = household. Prop. = proportion.

**Table S3:** Water contact per activity.

| Variable | Overall | Female | Male | P-value |
| --- | --- | --- | --- | --- |
| Getting drinking water (%) | 497 (17.3) | 348 (22.1) | 149 (11.5) | <0.001 |
| Washing clothes with soap (%) | 481 (16.8) | 368 (23.4) | 113 (8.7) | <0.001 |
| Fishing (%) | 354 (12.3) | 39 (2.5) | 315 (24.3) | <0.001 |
| Washing clothes without soap (%) | 152 (5.3) | 115 (7.3) | 37 (2.9) | <0.001 |
| Washing jerry cans or other household items (%) | 144 (5.0) | 108 (6.9) | 36 (2.8) | <0.001 |
| Fishmongering (%) | 104 (3.6) | 90 (5.7) | 14 (1.1) | <0.001 |
| Swimming or playing (%) | 78 (2.7) | 32 (2.0) | 46 (3.6) | 0.018 |
| Bathing with soap (%) | 49 (1.7) | 29 (1.8) | 20 (1.5) | 0.640 |
| Bathing without soap (%) | 38 (1.3) | 19 (1.2) | 19 (1.5) | 0.658 |
| Collecting papyrus (%) | 16 (0.6) | 8 (0.5) | 8 (0.6) | 0.888 |
| Collecting shells (%) | 15 (0.5) | 11 (0.7) | 4 (0.3) | 0.238 |

*Note:*
Number and percentage of participants with water contact [n (%)] per activity. Chi-squared tests were used to test for group differences.

**Table S4:**
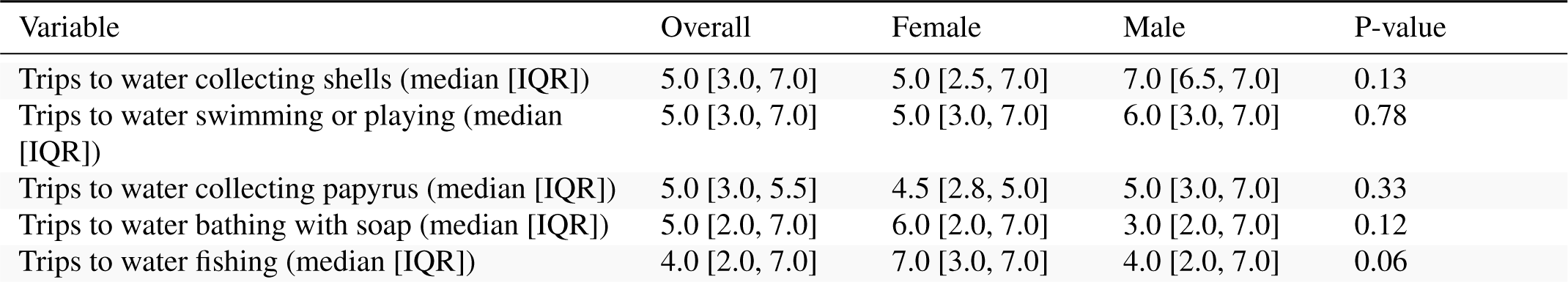
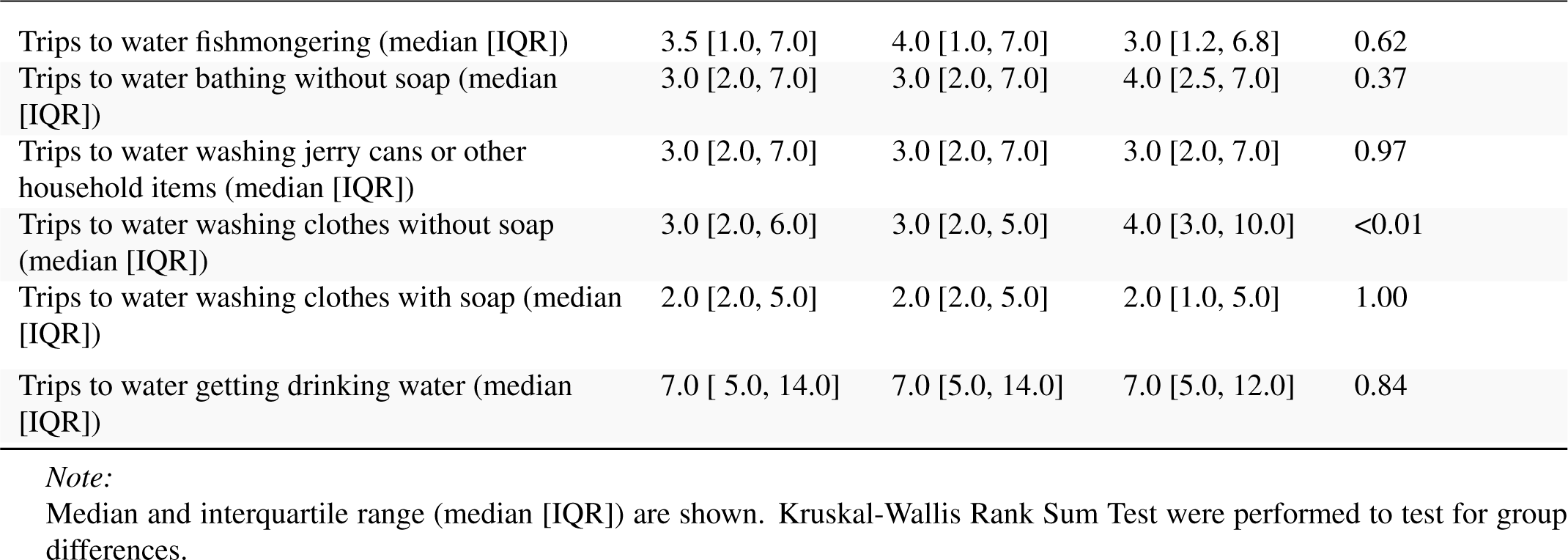
Water contact frequency per activity (in trips per week)

| Variable | Overall | Female | Male | P-value |
| --- | --- | --- | --- | --- |
| Trips to water collecting shells (median [IQR]) | 5.0 [3.0, 7.0] | 5.0 [2.5, 7.0] | 7.0 [6.5, 7.0] | 0.13 |
| Trips to water swimming or playing (median [IQR]) | 5.0 [3.0, 7.0] | 5.0 [3.0, 7.0] | 6.0 [3.0, 7.0] | 0.78 |
| Trips to water collecting papyrus (median [IQR]) | 5.0 [3.0, 5.5] | 4.5 [2.8, 5.0] | 5.0 [3.0, 7.0] | 0.33 |
| Trips to water bathing with soap (median [IQR]) | 5.0 [2.0, 7.0] | 6.0 [2.0, 7.0] | 3.0 [2.0, 7.0] | 0.12 |
| Trips to water fishing (median [IQR]) | 4.0 [2.0, 7.0] | 7.0 [3.0, 7.0] | 4.0 [2.0, 7.0] | 0.06 |
| Trips to water fishmongering (median [IQR]) | 3.5 [1.0, 7.0] | 4.0 [1.0, 7.0] | 3.0 [1.2, 6.8] | 0.62 |
| Trips to water bathing without soap (median [IQR]) | 3.0 [2.0, 7.0] | 3.0 [2.0, 7.0] | 4.0 [2.5, 7.0] | 0.37 |
| Trips to water washing jerry cans or other household items (median [IQR]) | 3.0 [2.0, 7.0] | 3.0 [2.0, 7.0] | 3.0 [2.0, 7.0] | 0.97 |
| Trips to water washing clothes without soap (median [IQR]) | 3.0 [2.0, 6.0] | 3.0 [2.0, 5.0] | 4.0 [3.0, 10.0] | <0.01 |
| Trips to water washing clothes with soap (median [IQR]) | 2.0 [2.0, 5.0] | 2.0 [2.0, 5.0] | 2.0 [1.0, 5.0] | 1.00 |
| Trips to water getting drinking water (median [IQR]) | 7.0 [ 5.0, 14.0] | 7.0 [5.0, 14.0] | 7.0 [5.0, 12.0] | 0.84 |

*Note:*
Median and interquartile range (median [IQR]) are shown. Kruskal-Wallis Rank Sum Test were performed to test for group differences.

**Table S5:** Water contact duration per activity (in hrs per week)

| Variable | Overall | Female | Male | P-value |
| --- | --- | --- | --- | --- |
| Time spent fishing (hrs) (median [IQR]) | 4.0 [3.0, 4.0] | 4.0 [2.5, 4.0] | 4.0 [3.0, 4.0] | 0.81 |
| Time spent collecting papyrus (hrs) (median [IQR]) | 3.0 [2.0, 3.0] | 3.0 [3.0, 4.0] | 2.0 [1.8, 3.0] | 0.01 |
| Time spent collecting shells (hrs) (median [IQR]) | 2.0 [2.0, 3.5] | 3.0 [2.0, 4.0] | 2.0 [2.0, 2.2] | 0.33 |
| Time spent fishmongering (hrs) (median [IQR]) | 2.0 [1.0, 3.0] | 2.0 [1.0, 3.0] | 2.0 [1.0, 3.0] | 0.98 |
| Time spent washing clothes without soap (hrs) (median [IQR]) | 2.0 [1.0, 3.0] | 2.0 [1.0, 3.0] | 2.0 [1.0, 2.0] | 0.90 |
| Time spent washing clothes with soap (hrs) (median [IQR]) | 2.0 [1.0, 2.0] | 2.0 [1.0, 2.0] | 2.0 [1.0, 3.0] | 0.61 |
| Time spent swimming or playing (hrs) (median [IQR]) | 1.0 [1.0, 2.0] | 1.0 [0.5, 3.0] | 1.0 [1.0, 2.0] | 0.92 |
| Time spent bathing without soap (hrs) (median [IQR]) | 1.0 [0.5, 2.0] | 1.0 [0.8, 2.0] | 1.0 [0.5, 2.0] | 0.49 |
| Time spent washing jerry cans or other household items (hrs) (median [IQR]) | 1.0 [0.5, 1.2] | 1.0 [0.5, 1.2] | 1.0 [1.0, 1.2] | 0.53 |
| Time spent getting drinking water (hrs) (median [IQR]) | 1.0 [0.5, 1.0] | 1.0 [0.5, 1.0] | 1.0 [0.5, 1.0] | 0.54 |
| Time spent bathing with soap (hrs) (median [IQR]) | 0.5 [0.5, 2.0] | 0.5 [0.5, 2.0] | 0.5 [0.5, 2.0] | 0.90 |

*Note:*
Median and interquartile range (median [IQR]) are shown. Kruskal-Wallis Rank Sum Test were performed to test for group differences.

**Table S6:**
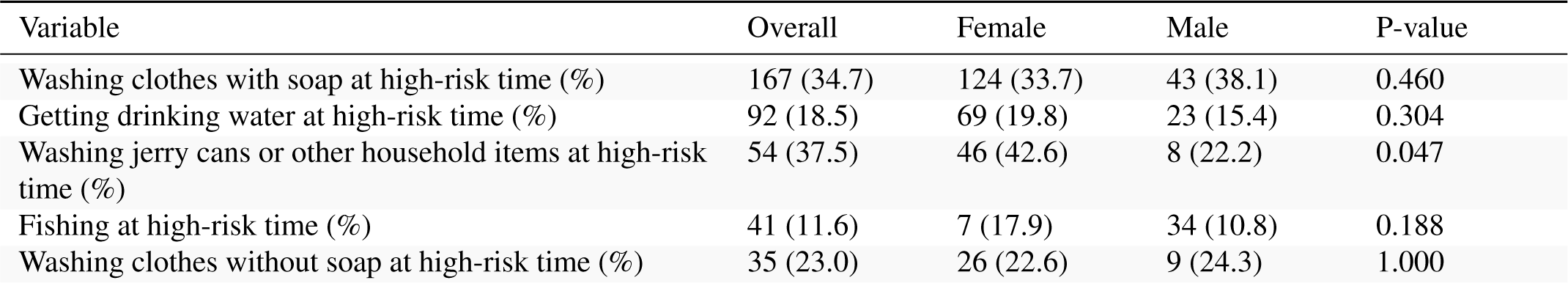
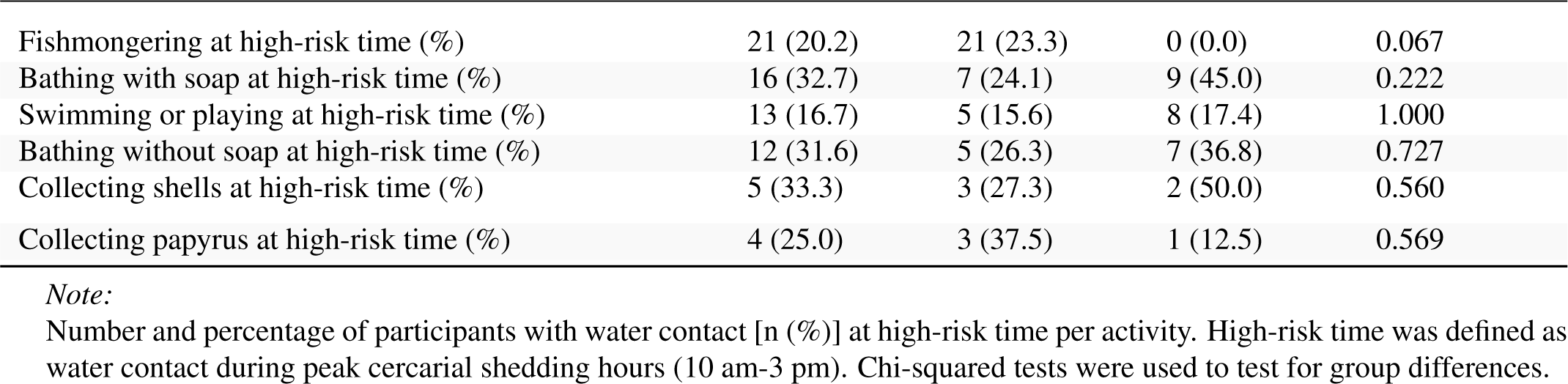
Water contact at high-risk time per activity.

| Variable | Overall | Female | Male | P-value |
| --- | --- | --- | --- | --- |
| Washing clothes with soap at high-risk time (%) | 167 (34.7) | 124 (33.7) | 43 (38.1) | 0.460 |
| Getting drinking water at high-risk time (%) | 92 (18.5) | 69 (19.8) | 23 (15.4) | 0.304 |
| Washing jerry cans or other household items at high-risk time (%) | 54 (37.5) | 46 (42.6) | 8 (22.2) | 0.047 |
| Fishing at high-risk time (%) | 41 (11.6) | 7 (17.9) | 34 (10.8) | 0.188 |
| Washing clothes without soap at high-risk time (%) | 35 (23.0) | 26 (22.6) | 9 (24.3) | 1.000 |
| Fishmongering at high-risk time (%) | 21 (20.2) | 21 (23.3) | 0 (0.0) | 0.067 |
| Bathing with soap at high-risk time (%) | 16 (32.7) | 7 (24.1) | 9 (45.0) | 0.222 |
| Swimming or playing at high-risk time (%) | 13 (16.7) | 5 (15.6) | 8 (17.4) | 1.000 |
| Bathing without soap at high-risk time (%) | 12 (31.6) | 5 (26.3) | 7 (36.8) | 0.727 |
| Collecting shells at high-risk time (%) | 5 (33.3) | 3 (27.3) | 2 (50.0) | 0.560 |
| Collecting papyrus at high-risk time (%) | 4 (25.0) | 3 (37.5) | 1 (12.5) | 0.569 |

*Note:*
Number and percentage of participants with water contact [n (%)] at high-risk time per activity. High-risk time was defined as water contact during peak cercarial shedding hours (10 am-3 pm). Chi-squared tests were used to test for group differences.

**Table S7:**
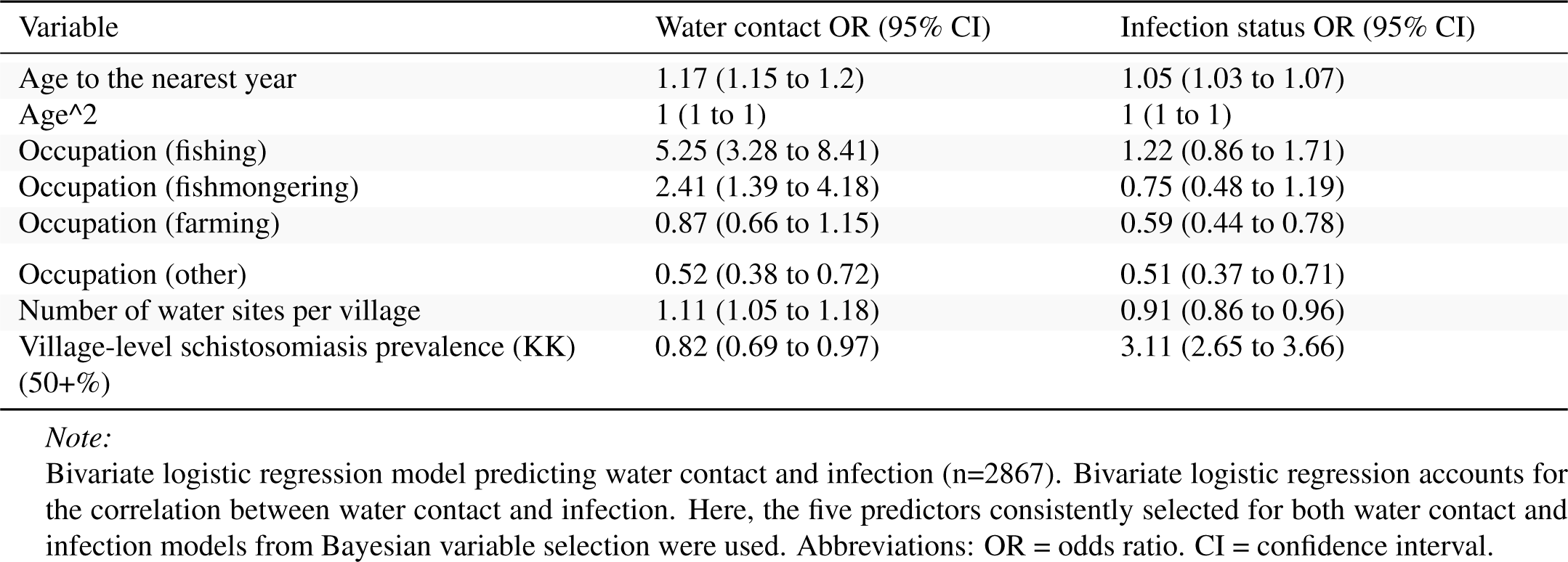
Bivariate logistic model (water contact and infection status)

| Variable | Water contact OR (95% CI) | Infection status OR (95% CI) |
| --- | --- | --- |
| Age to the nearest year | 1.17 (1.15 to 1.2) | 1.05 (1.03 to 1.07) |
| Age<sup>2</sup> | 1 (1 to 1) | 1 (1 to 1) |
| Occupation (fishing) | 5.25 (3.28 to 8.41) | 1.22 (0.86 to 1.71) |
| Occupation (fishmongering) | 2.41 (1.39 to 4.18) | 0.75 (0.48 to 1.19) |
| Occupation (farming) | 0.87 (0.66 to 1.15) | 0.59 (0.44 to 0.78) |
| Occupation (other) | 0.52 (0.38 to 0.72) | 0.51 (0.37 to 0.71) |
| Number of water sites per village | 1.11 (1.05 to 1.18) | 0.91 (0.86 to 0.96) |
| Village-level schistosomiasis prevalence (KK) (50+%) | 0.82 (0.69 to 0.97) | 3.11 (2.65 to 3.66) |

*Note:*
Bivariate logistic regression model predicting water contact and infection (n=2867). Bivariate logistic regression accounts for the correlation between water contact and infection. Here, the five predictors consistently selected for both water contact and infection models from Bayesian variable selection were used. Abbreviations: OR = odds ratio, CI = confidence interval.

**Table S8:** Comparison of variable selection between LRTs and BVS.

| variable | Infection status (LRT) | Infection status (BAS) | Water contact (LRT) | Water contact (BAS) |
| --- | --- | --- | --- | --- |
| Age to the nearest year | X | X | X | X |
| Age<sup>2</sup> | X | X | X | X |
| Female | X | - | X | X |
| Level of education | X | X | X | - |
| Enrolled at school | X | - | X | - |
| Occupational water contact | X | - | - | - |
| Number of water contact activities | X | - | - | - |
| Occupation | X | X | X | X |
| Contamination at water site closest to HH | - | - | X | X |
| Closest water site to HH in village | - | - | X | - |
| HH has handwashing facility with soap | X | - | - | - |
| Home latrine | X | - | - | - |

| variable | Infection<br>status<br>(LRT) | Infection<br>status<br>(BAS) | Water<br>contact<br>(LRT) | Water<br>contact<br>(BAS) |
| --- | --- | --- | --- | --- |
| Prop. of HHs with safe drinking water per village | - | - | - | X |
| Distance to closest water site from HH (km) | X | - | X | - |
| Distance to closest water site from village centre (km) | - | - | X | - |
| Water site type in village | - | X | - | - |
| Type of water site closest to HH | X | - | - | X |
| Number of water sites per village | - | X | - | X |
| Village-level schistosomiasis prevalence (KK) | - | X | - | X |
| Number of roads per village | - | - | - | X |

*Note:*
Comparison of variable sets selected using likelihood ratio tests (LRTs) at $p < 0.05$ with the variable sets selected via Bayesian variable selection (BVS) with marginal inclusion probabilities $p \geq 0.5$ (i.e., variables included in the median probability model).

## 2 Supplementary Figures

**Figure S1:**
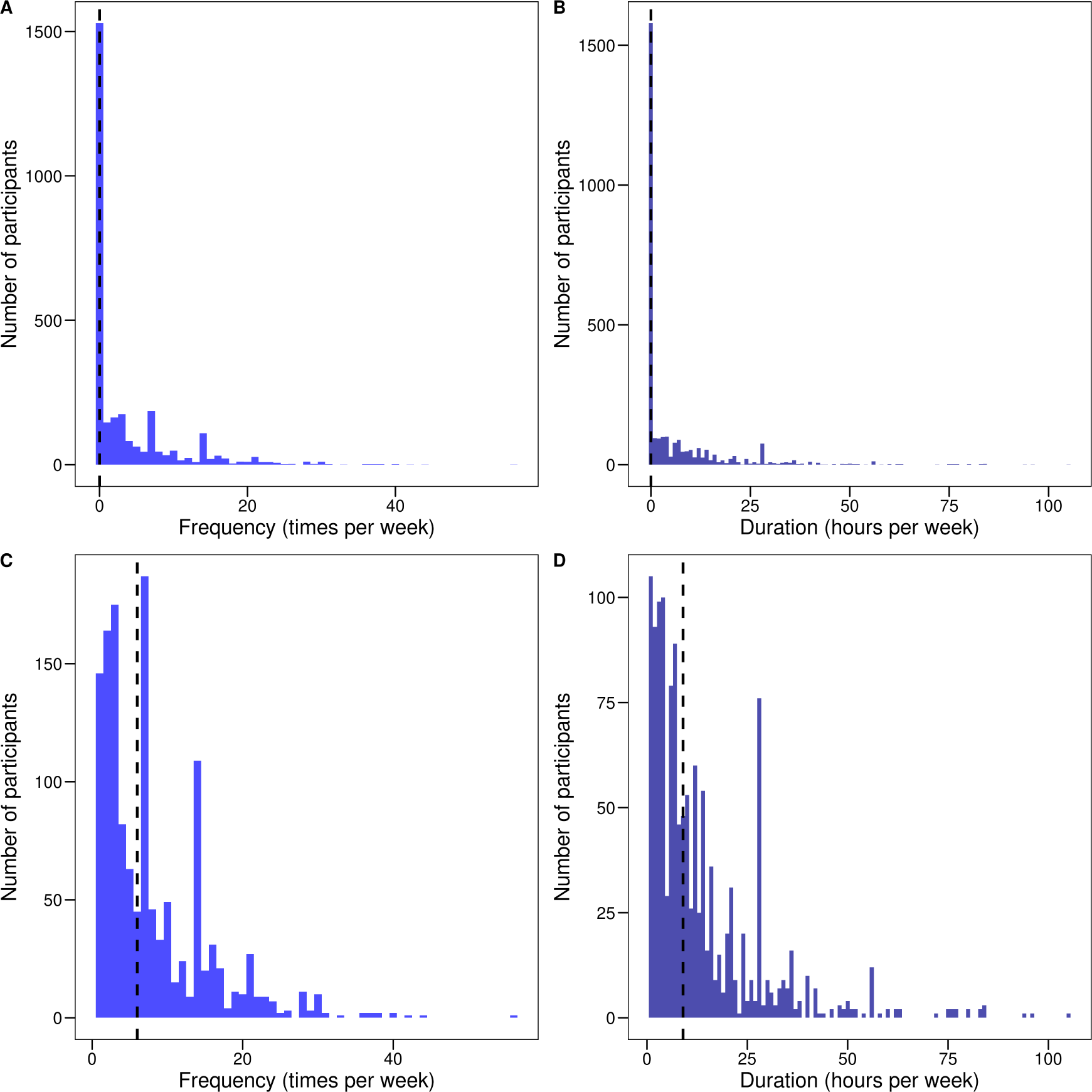
**A.** Distribution of water contact frequency among all participants. **B.** Distribution of water contact duration among all participants. **C.** Distribution of water contact frequency among participants with water contact only. **D.** Distribution of water contact duration among participants with water contact only. Dashed vertical line = median.

**Figure S2:**
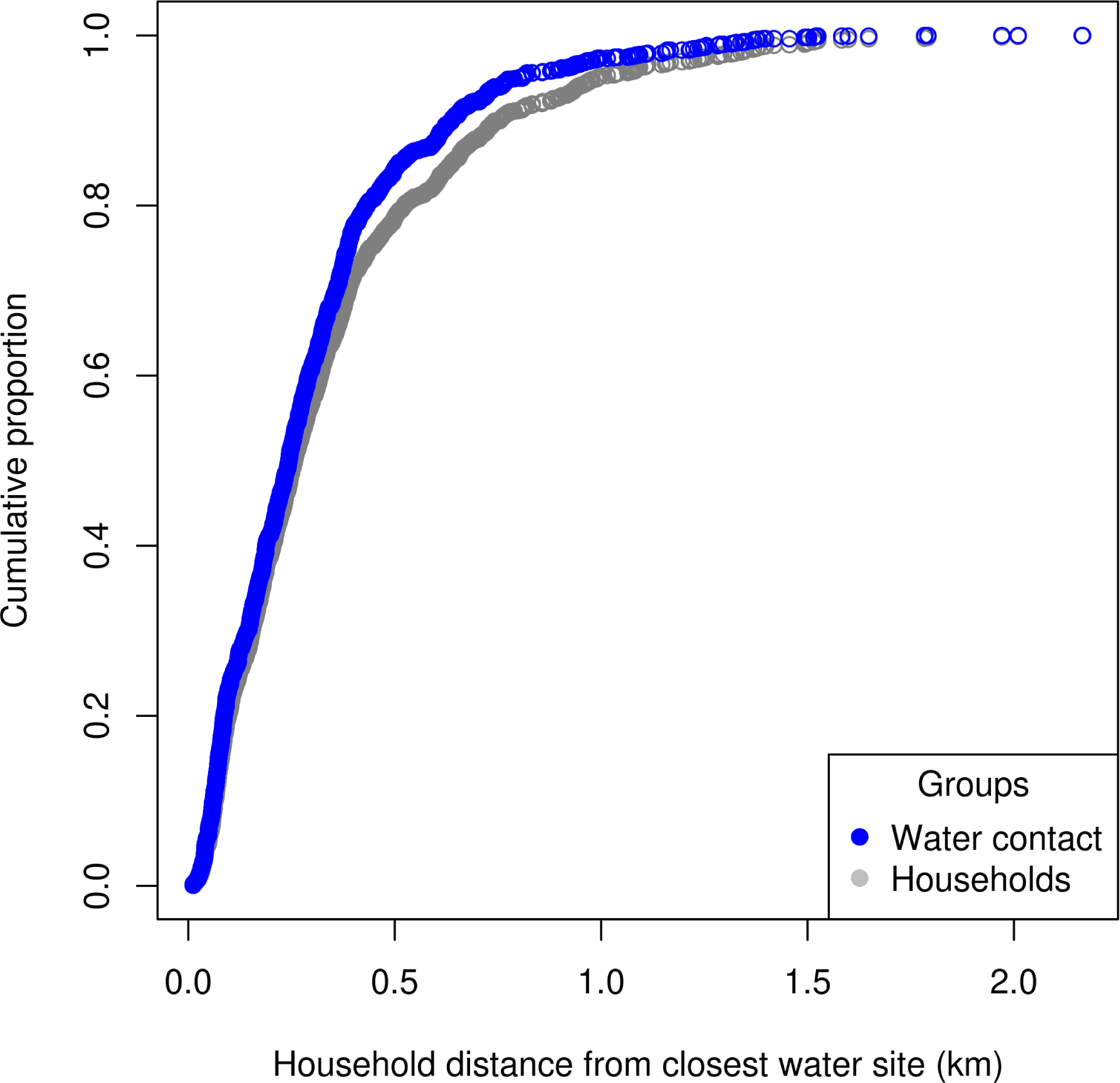
Cumulative proportion of all water contacts/participants by household distance to the closest water site (n=2867).

**Figure S3:**
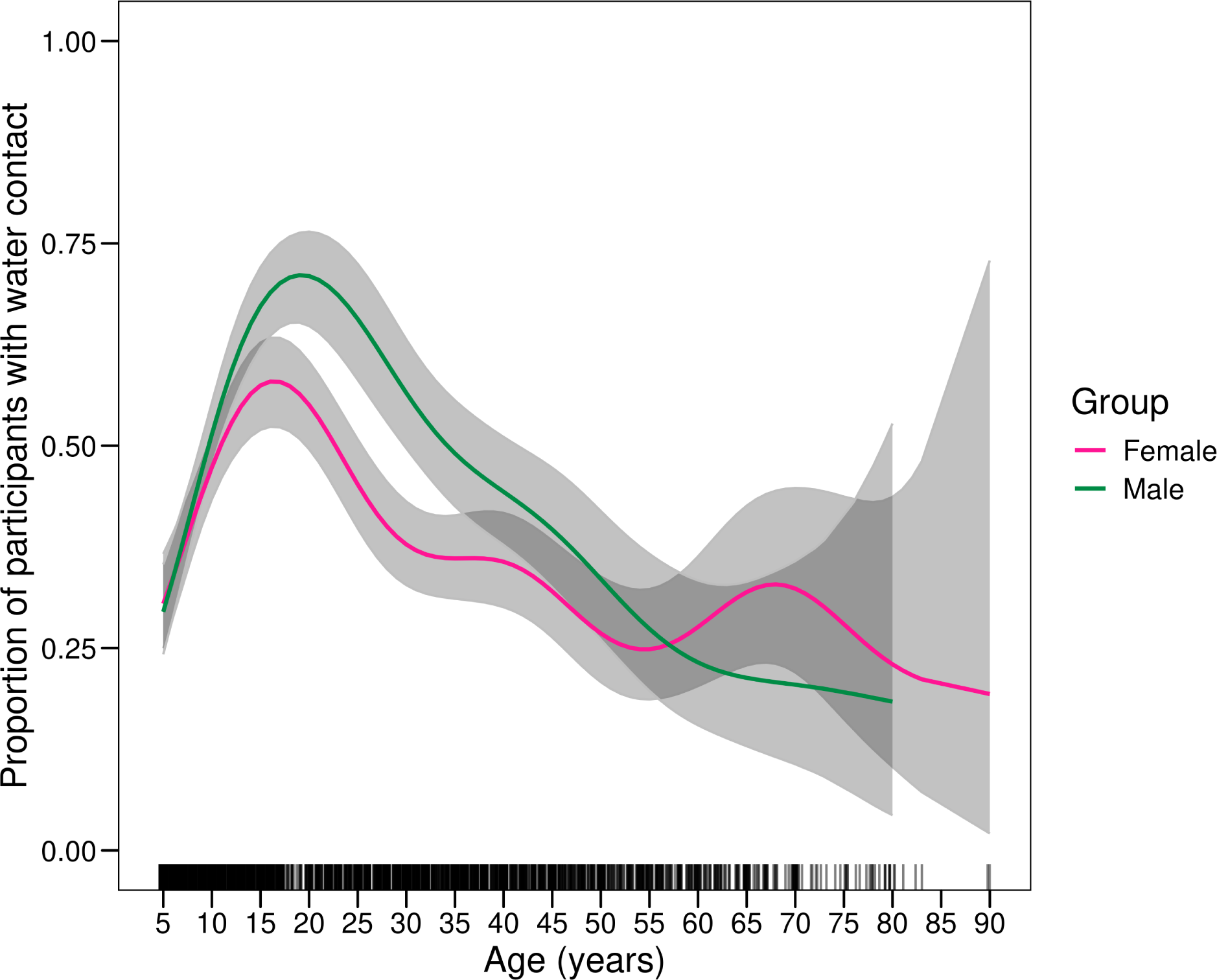
Generalised additive model predicting the proportion of participants with water contact over age and gender.

**Figure S4:**
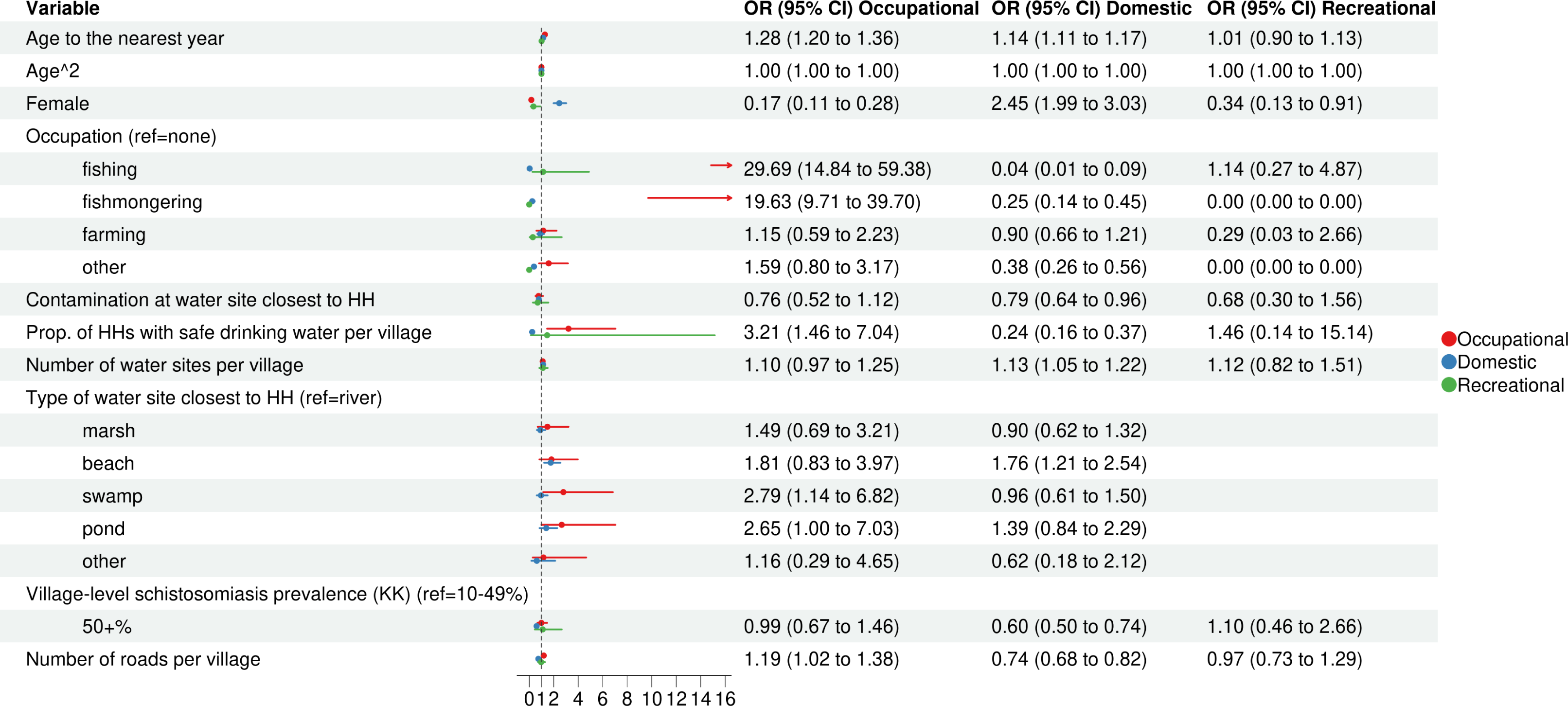
Results from seperate logistic regression models with standard errors clustered at the household level predicting domestic, occupational, and recreational water contact among participants that do not engage in multiple types of water contact (n=2,684). As not all categories in the water site type variable were sufficiently represented in the recreational water contact model, we removed this variable when predicting recreational water contact. Abbreviations: OR = odds ratio. CI = confidence interval.

**Figure S5:**
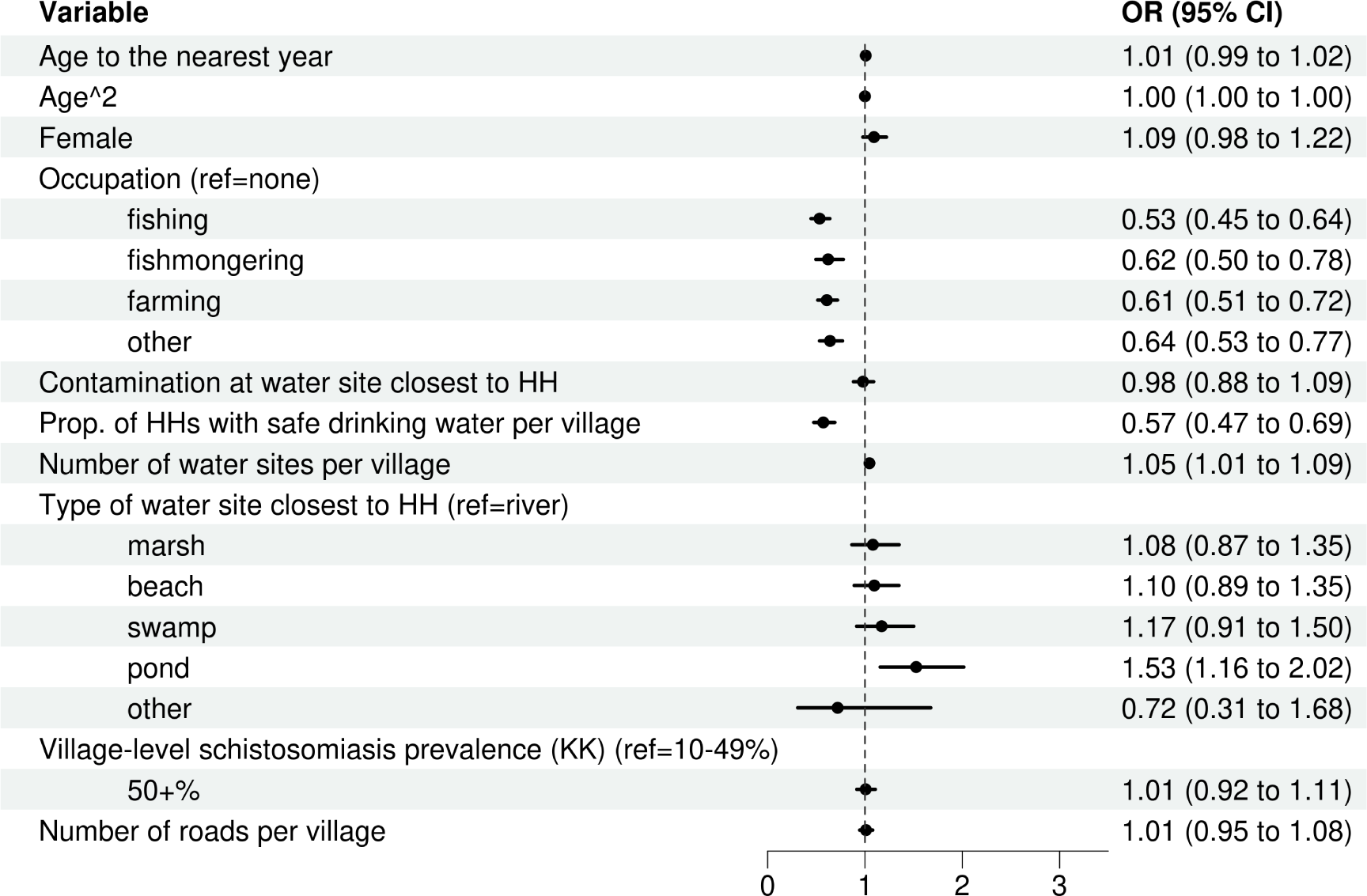
Negative binomial regression model predicting water contact frequency among participants with water contact (n=1,339). Standard errors clustered at the household level. Abbreviations: OR = odds ratio. CI = confidence interval.

**Figure S6:**
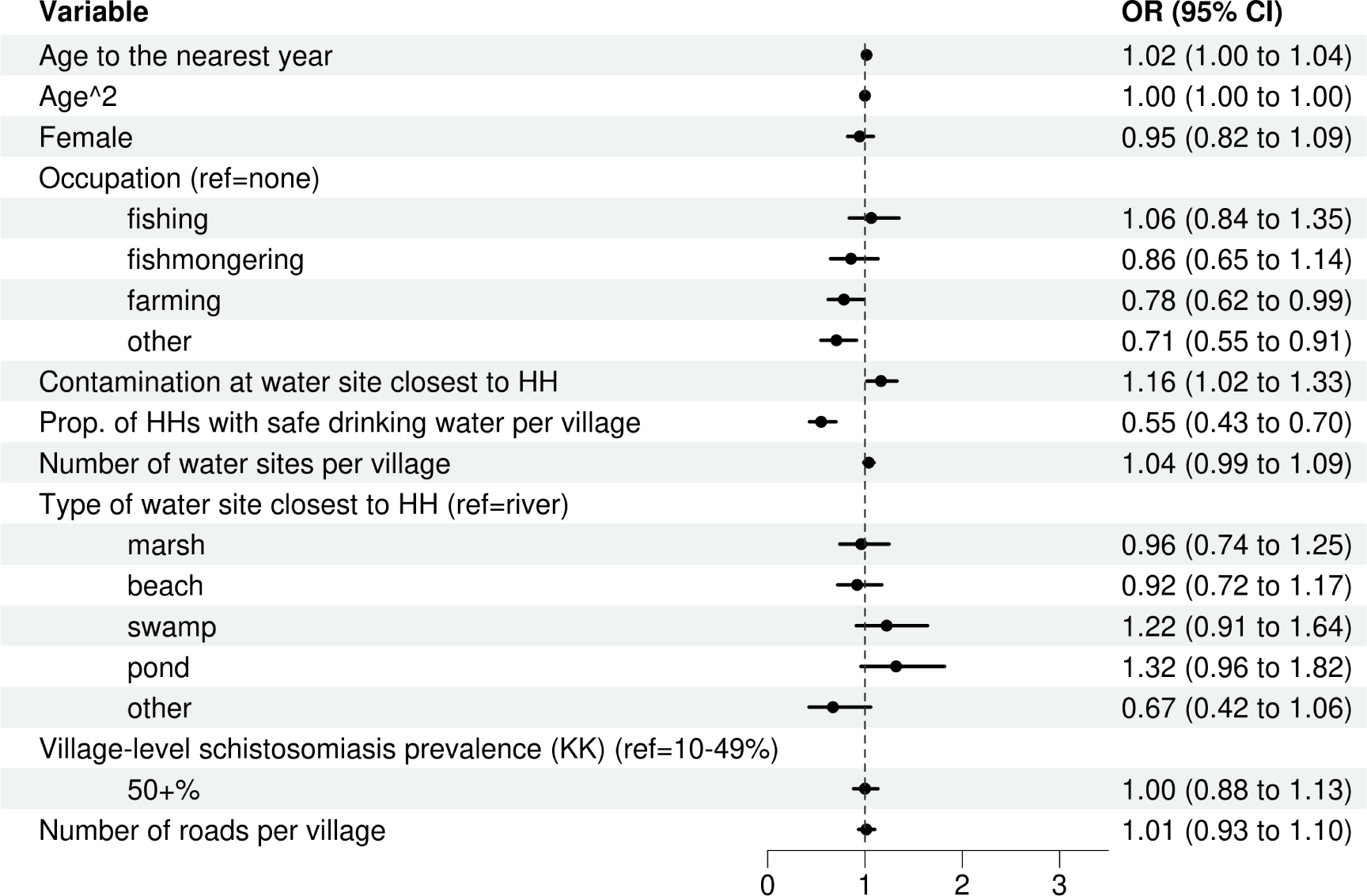
Negative binomial regression model predicting water contact duration among participants with water contact (n=1,339). Standard errors clustered at the household level. Abbreviations: OR = odds ratio. CI = confidence interval.

**Figure S7:**
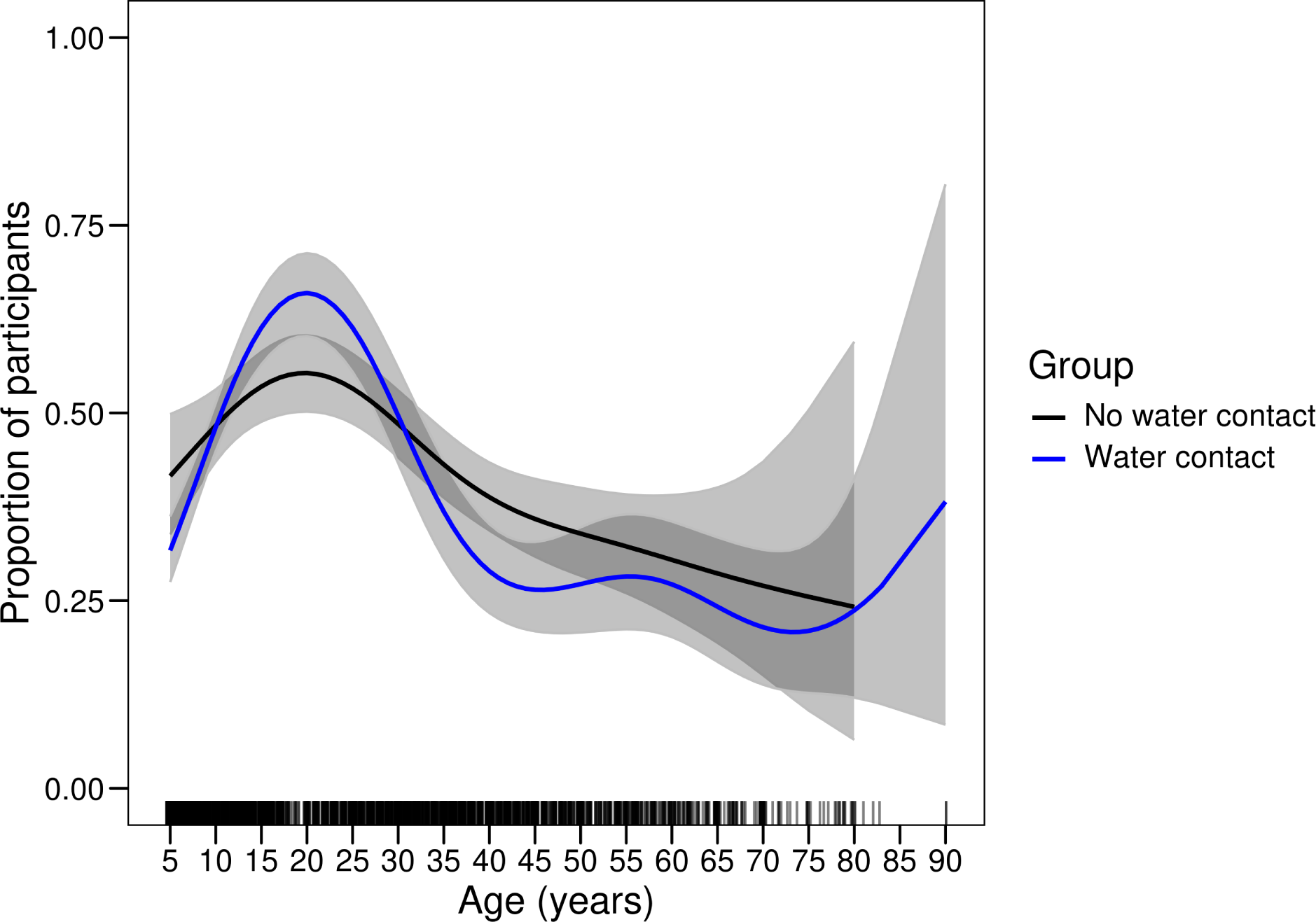
Generalised additive model predicting the proportion of participants with *S. mansoni* infection and heavy infection (400+ eggs per gram of stool, by Kato-Katz micrsocopy) as well as the proportion of participants with water contact over household distance to the closest water site (n=2867).

**Figure S8:**
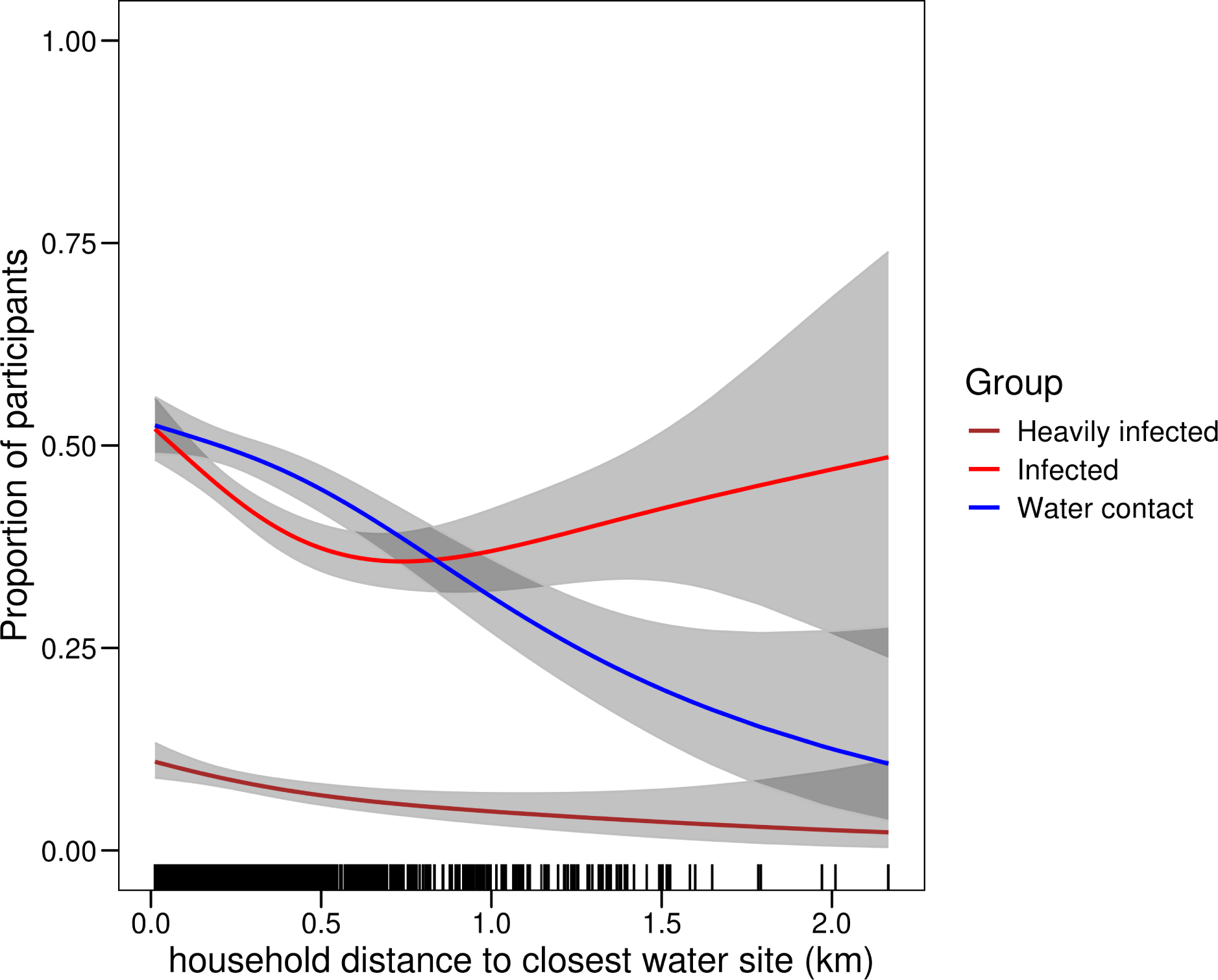
Generalised additive model predicting the proportion of participants with *S. mansoni* infection over water contact and age (i.e., comparing infection outcomes in participants with current water contact vs participants without current water contact over age, n=2867).

**Figure S9:**
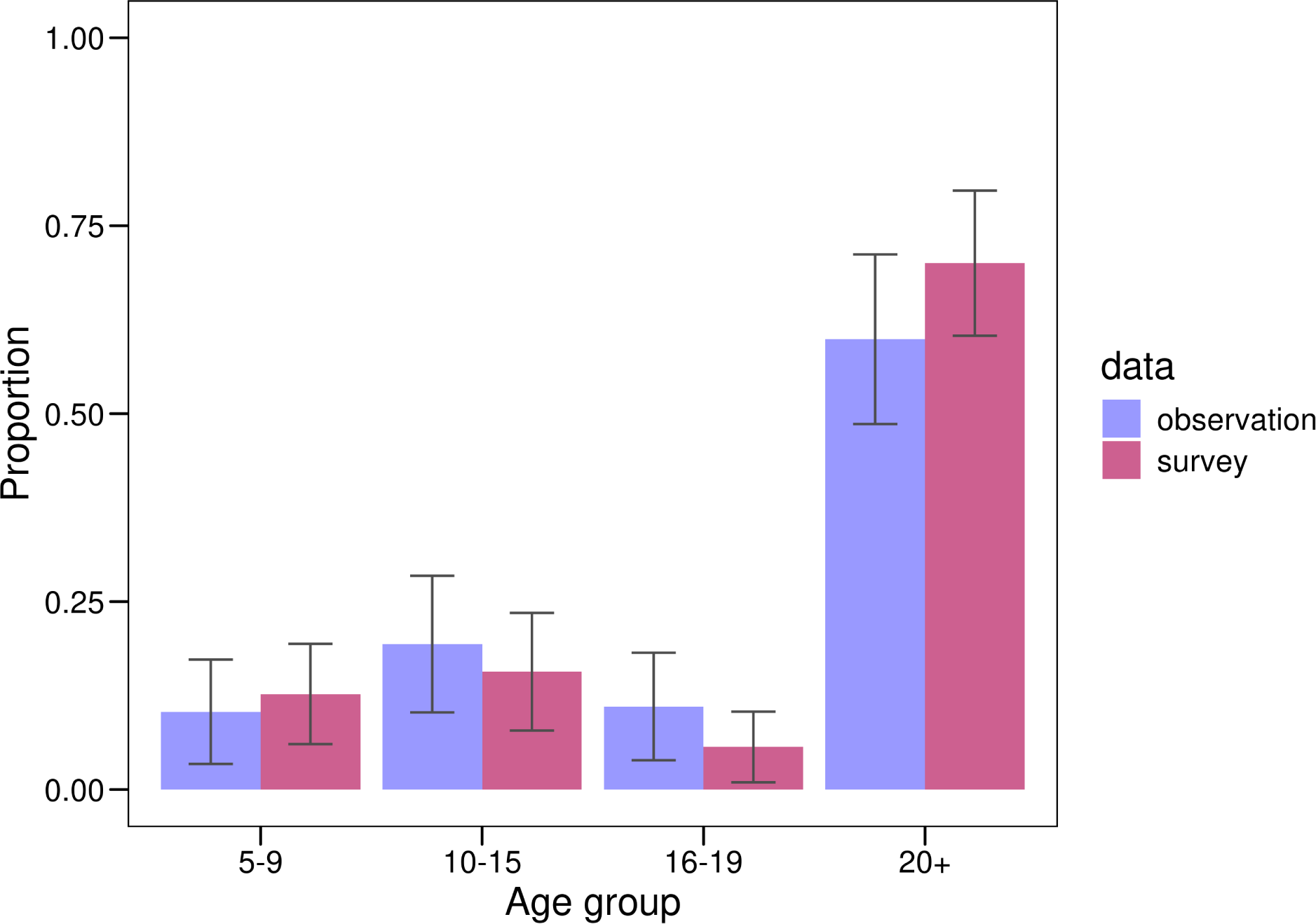
Comparison of age distribution in self-reported data with age distribution of direct observation data in 12 villages with both observation and survey data (survey data n=605 and direct observation n=13,515). Proportions represent the proportion of all water contacts which occur in each age group. Age groups were based on the categories used for MDA treatment and used because direct observation was conducted using these age groups. Whiskers represent 95% confidence intervals obtained using bootstrapping with 1000 repeats.

**Figure S10:**
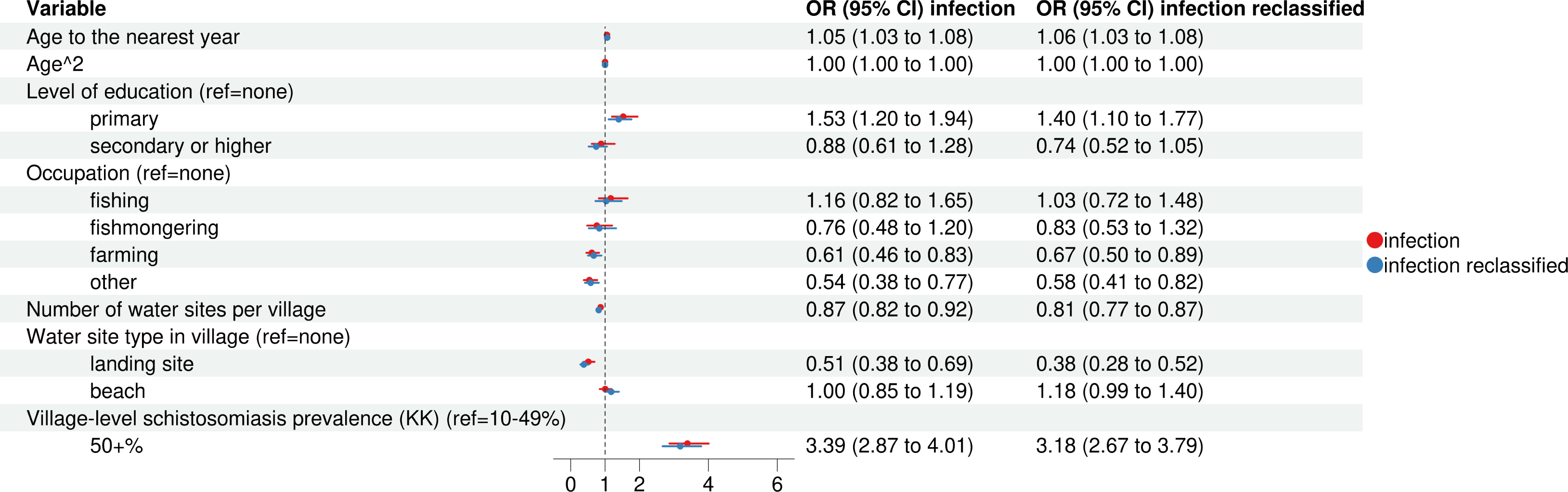
Seperate logistic regression model predicting infection status based on Kato-Katz as well as reclassified Kato-Katz (where participants were recoded as infected whenever Kato-Katz infection status was negative but when the more sensitive POC-CCA diagnostic (positive band 1-3) indicated an infection, n=2867). Standard errors clustered at the household level. Abbreviations: OR = odds ratio. CI = confidence interval.

**Figure S11:**
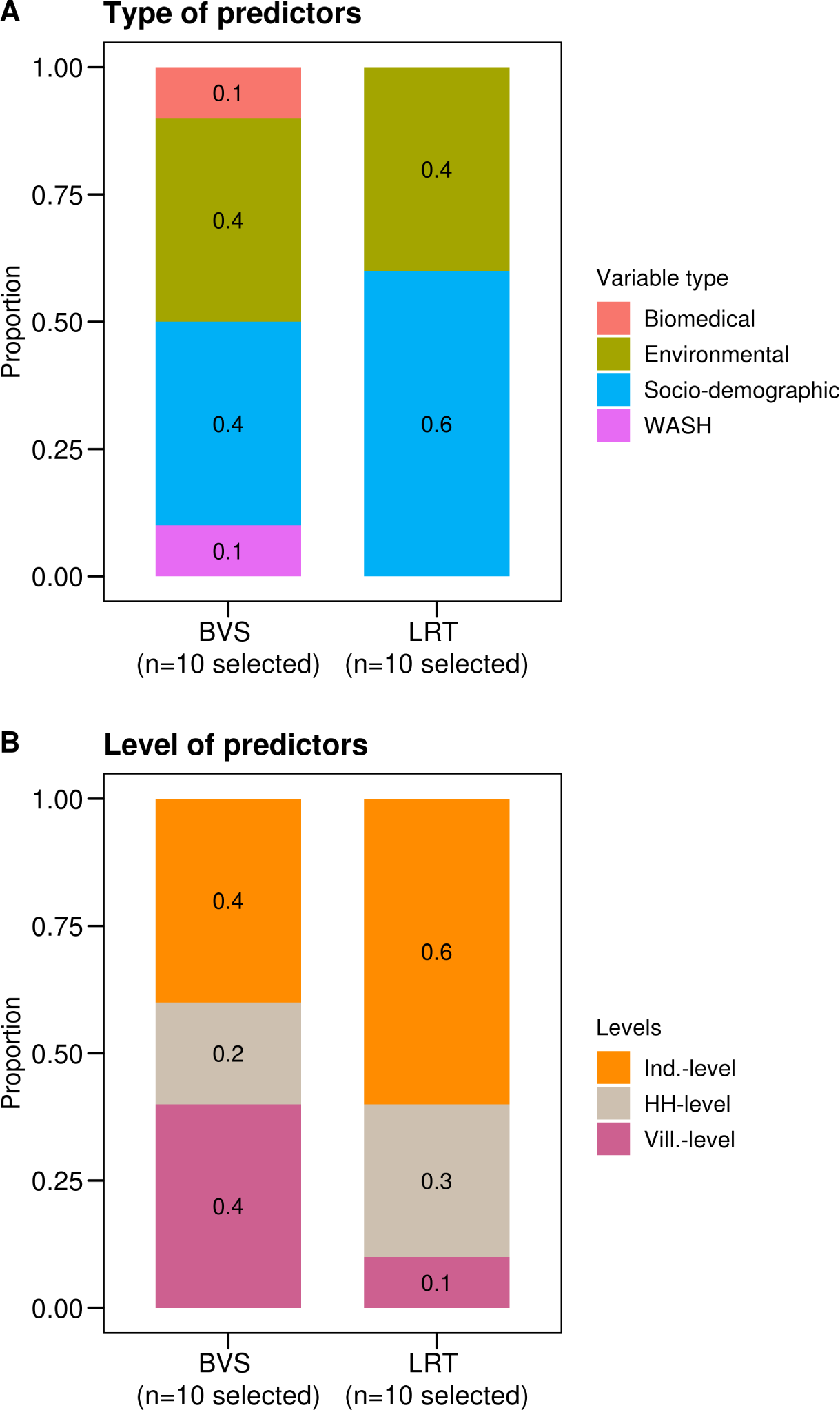
Comparison of variable selection results from BVS and LRTs for predicting water contact. **A**. shows the percentage of selected variables from Bayesian variable selection (BVS, n=10 selected variables) and likelihood ratio tests (LRTs, n=10 selected variables) by type of variable. Variables selected using LRTs were solely socio-demographic and environmental variables, while BAS also selected water, sanitation, and hygiene (WASH) and biomedical variables. Among socio-demographic variables, age, age^2^, gender, and occupation were consistently selected by BAS and LRTs. School enrolment status and level of education were only selected by LRT. Among environmental variables, contamination at the closest water site, and the number of roads per village were consistently selected. Distance variables were only selected by LRTs (household distance to nearest water site and village centre distance). BVS selected the number of water sites per village, the type of site closest to the household, contamination at nearest site, and village-level infection prevalence. No WASH variables were selected via LRT. In BVS, the WASH variables number of latrines per village and proportion of households using an improved drinking water source were selected. **B**. compares selected variables from BVS and LRTs by level. In LRTs, 60% of selected variables were individual level, while the same figure was 40% for BAS. The proportion of village-level variables was four times as high in the BVS variable set (40%) compared to the set from LRTs (10%).

**Figure S12:**
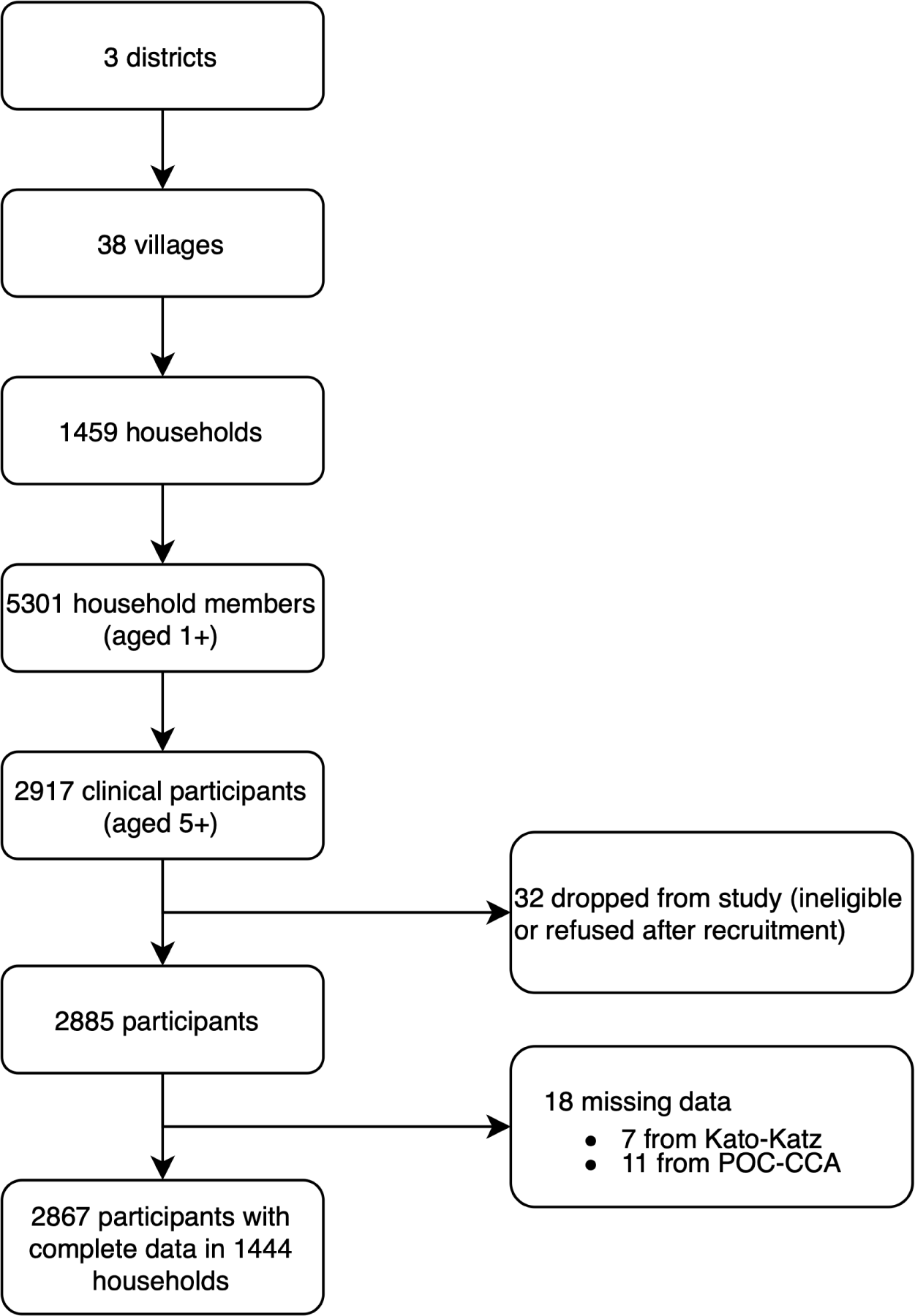
Participant flow diagram

## Notes

### Competing Interest Statement

The authors have declared no competing interest.

## References

1. Sokolow SH, Nova N, Jones IJ, Wood CL, Lafferty KD, Garchitorena A, et al. Ecological and socioeconomic factors associated with the human burden of environmentally mediated pathogens: a global analysis. Lancet Planet Health. 2022;6: e870–e879. doi:10.1016/S2542-5196(22)00248-0

2. Lai Y-S, Biedermann P, Ekpo UF, Garba A, Mathieu E, Midzi N, et al. Spatial distribution of schistosomiasis and treatment needs in sub-Saharan Africa: a systematic review and geostatistical analysis. Lancet Infect Dis. 2015;15: 927–940. doi:10.1016/S1473-3099(15)00066-3

3. Reitzug F, Ledien J, Chami GF. Associations of water contact frequency, duration, and activities with schistosome infection risk: A systematic review and meta-analysis. Freeman MC, editor. PLoS Negl Trop Dis. 2023;17: e0011377. doi:10.1371/journal.pntd.0011377

4. World Health Organization. WHO guideline on control and elimination of human schistosomiasis. Geneva: World Health Organization; 2022. Available: https://apps.who.int/iris/handle/10665/351856

5. Chami GF, Kontoleon AA, Bulte E, Fenwick A, Kabatereine NB, Tukahebwa EM, et al. Profiling Nonrecipients of Mass Drug Administration for Schistosomiasis and Hookworm Infections: A Comprehensive Analysis of Praziquantel and Albendazole Coverage in Community-Directed Treatment in Uganda. Clin Infect Dis. 2016;62: 200–207. doi:10.1093/cid/civ829

6. Crellen T, Walker M, Lamberton PHL, Kabatereine NB, Tukahebwa EM, Cotton JA, et al. Reduced Efficacy of Praziquantel Against *Schistosoma mansoni* Is Associated With Multiple Rounds of Mass Drug Administration. Clin Infect Dis. 2016; ciw506. doi:10.1093/cid/ciw506

7. World Health Organization. Ending the neglect to attain the Sustainable Development Goals: a road map for neglected tropical diseases 2021–2030. 2020.

8. Hoover CM, Sokolow SH, Kemp J, Sanchirico JN, Lund AJ, Jones IJ, et al. Modelled effects of prawn aquaculture on poverty alleviation and schistosomiasis control. Nat Sustain. 2019;2: 611–620. doi:10.1038/s41893-019-0301-7

9. Knopp S, Person B, Ame SM, Ali SM, Hattendorf J, Juma S, et al. Evaluation of integrated interventions layered on mass drug administration for urogenital schistosomiasis elimination: a cluster-randomised trial. Lancet Glob Health. 2019;7: e1118–e1129. doi:10.1016/S2214-109X(19)30189-5

10. Torres-Vitolas CA, Trienekens SCM, Zaadnoordijk W, Gouvras AN. Behaviour change interventions for the control and elimination of schistosomiasis: A systematic review of evidence from low- and middle-income countries. Ramos AN, editor. PLoS Negl Trop Dis. 2023;17: e0011315. doi:10.1371/journal.pntd.0011315

11. Rohr JR, Sack A, Bakhoum S, Barrett CB, Lopez-Carr D, Chamberlin AJ, et al. A planetary health innovation for disease, food and water challenges in Africa. Nature. 2023;619: 782–787. doi:10.1038/s41586-023-06313-z

12. Grimes JET, Croll D, Harrison WE, Utzinger J, Freeman MC, Templeton MR. The Relationship between Water, Sanitation and Schistosomiasis: A Systematic Review and Meta-analysis. McGarvey ST, editor. PLoS Negl Trop Dis. 2014;8: e3296. doi:10.1371/journal.pntd.0003296

13. Scott J, Diakhate M, Vereecken K, Fall A, Diop M, Ly A, et al. Human water contacts patterns in *Schistosoma mansoni* epidemic foci in northern Senegal change according to age, sex and place of residence, but are not related to intensity of infection. Trop Med Int Health. 2003;8: 100–108. doi:10.1046/j.1365-3156.2003.00993.x

14. Rudge JW, Stothard JR, Basáñez M-G, Mgeni AF, Khamis IS, Khamis AN, et al. Micro-epidemiology of urinary schistosomiasis in Zanzibar: Local risk factors associated with distribution of infections among schoolchildren and relevance for control. Acta Trop. 2008;105: 45–54. doi:10.1016/j.actatropica.2007.09.006

15. Lund AJ, Sokolow SH, Jones IJ, Wood CL, Ali S, Chamberlin A, et al. Exposure, hazard, and vulnerability all contribute to *Schistosoma haematobium* re-infection in northern Senegal. Lamberton PHL, editor. PLoS Negl Trop Dis. 2021;15: e0009806. doi:10.1371/journal.pntd.0009806

16. Etard J-F, Audibert M, Dabo A. Age-Acquired Resistance and Predisposition to Reinfection with *Schistosoma haematobium* after Treatment with Praziquantel in Mali. Am J Trop Med Hyg. 1995;52: 549–558. doi:10.4269/ajtmh.1995.52.549

17. Pearce EJ, MacDonald AS. The immunobiology of schistosomiasis. Nat Rev Immunol. 2002;2: 499–511. doi:10.1038/nri843

18. Joseph S, Jones FM, Walter K, Fulford AJ, Kimani G, Mwatha JosephK, et al. Increases in Human T Helper 2 Cytokine Responses to *Schistosoma mansoni* Worm and WormLTegument Antigens Are Induced by Treatment with Praziquantel. J Infect Dis. 2004;190: 835–842. doi:10.1086/422604

19. Joseph S, Jones FM, Kimani G, Mwatha JK, Kamau T, Kazibwe F, et al. Cytokine Production in Whole Blood Cultures from a Fishing Community in an Area of High Endemicity for *Schistosoma mansoni* in Uganda: The Differential Effect of Parasite Worm and Egg Antigens. Infect Immun. 2004;72: 728–734. doi:10.1128/IAI.72.2.728-734.2004

20. Arnold BF, Kanyi H, Njenga SM, Rawago FO, Priest JW, Secor WE, et al. Fine-scale heterogeneity in *Schistosoma mansoni* force of infection measured through antibody response. Proc Natl Acad Sci. 2020;117: 23174–23181. doi:10.1073/pnas.2008951117

21. Lamberti O, Kabatereine NB, Tukahebwa EM, Chami GF. *Schistosoma mansoni* infection risk for school-aged children clusters within households and is modified by distance to freshwater bodies. PLoS ONE. 2021;16: e0258915. doi:10.1371/journal.pone.0258915

22. Graham M, Ayabina D, Lucas TCD, Collyer BS, Medley GF, Hollingsworth TD, et al. SCHISTOX: An individual based model for the epidemiology and control of schistosomiasis. Infect Dis Model. 2021;6: 438–447. doi:10.1016/j.idm.2021.01.010

23. Fulford AJC, Butterworth AE, Ouma JH, Sturrock RF. A statistical approach to schistosome population dynamics and estimation of the life-span of *Schistosoma mansoni* in man. Parasitology. 1995;110: 307–316. doi:10.1017/S0031182000080896

24. Nuffield Department of Population Health, University of Oxford. SchistoTrack: a prospective multimorbidity cohort. [cited 16 Apr 2024]. Available: https://www.bdi.ox.ac.uk/research/schistotrack

25. Lo NC, Bezerra FSM, Colley DG, Fleming FM, Homeida M, Kabatereine N, et al. Review of 2022 WHO guidelines on the control and elimination of schistosomiasis. Lancet Infect Dis. 2022; S1473309922002213. doi:10.1016/S1473-3099(22)00221-3

26. Lo NC, Addiss DG, Hotez PJ, King CH, Stothard JR, Evans DS, et al. A call to strengthen the global strategy against schistosomiasis and soil-transmitted helminthiasis: the time is now. Lancet Infect Dis. 2017;17: e64–e69. doi:10.1016/S1473-3099(16)30535-7

27. Anderson RM, Turner HC, Farrell SH, Truscott JE. Chapter Four - Studies of the Transmission Dynamics, Mathematical Model Development and the Control of Schistosome Parasites by Mass Drug Administration in Human Communities. In: Basáñez MG, Anderson RM, editors. Advances in Parasitology. Academic Press; 2016. pp. 199–246. doi:10.1016/bs.apar.2016.06.003

28. Sandoval N, Siles-Lucas M, Aban JL, Pérez-Arellano JL, Gárate T, Muro A. *Schistosoma mansoni*: A diagnostic approach to detect acute schistosomiasis infection in a murine model by PCR. Exp Parasitol. 2006;114: 84–88. doi:10.1016/j.exppara.2006.02.012

29. McManus DP, Dunne DW, Sacko M, Utzinger J, Vennervald BJ, Zhou X-N. Schistosomiasis. Nat Rev Dis Primer. 2018;4: 13. doi:10.1038/s41572-018-0013-8

30. Ayabina DV, Clark J, Bayley H, Lamberton PHL, Toor J, Hollingsworth TD. Gender-related differences in prevalence, intensity and associated risk factors of *Schistosoma* infections in Africa: A systematic review and meta-analysis. Santos VS, editor. PLoS Negl Trop Dis. 2021;15: e0009083. doi:10.1371/journal.pntd.0009083

31. Freeman MC, Clasen T, Brooker SJ, Akoko DO, Rheingans R. The Impact of a School-Based Hygiene, Water Quality and Sanitation Intervention on Soil-Transmitted Helminth Reinfection: A Cluster-Randomized Trial. Am J Trop Med Hyg. 2013;89: 875–883. doi:10.4269/ajtmh.13-0237

32. Kosinski K, Adjei M, Bosompem K, Crocker J, Durant J, Osabutey D, et al. Effective Control of *Schistosoma haematobium* Infection in a Ghanaian Community following Installation of a Water Recreation Area. PLoS Negl Trop Dis. 2012;6. doi:10.1371/journal.pntd.0001709

33. Grimes JE, Croll D, Harrison WE, Utzinger J, Freeman MC, Templeton MR. The roles of water, sanitation and hygiene in reducing schistosomiasis: a review. Parasit Vectors. 2015;8: 156. doi:10.1186/s13071-015-0766-9

34. Wiegand RE, Mwinzi PNM, Montgomery SP, Chan YL, Andiego K, Omedo M, et al. A Persistent Hotspot of *Schistosoma mansoni* Infection in a Five-Year Randomized Trial of Praziquantel Preventative Chemotherapy Strategies. J Infect Dis. 2017;216: 1425– 1433. doi:10.1093/infdis/jix496

35. Kittur N, Campbell CH, Binder S, Shen Y, Wiegand RE, Mwanga JR, et al. Discovering, Defining, and Summarizing Persistent Hotspots in SCORE Studies. Am J Trop Med Hyg. 2020;103: 24–29. doi:10.4269/ajtmh.19-0815

36. Amoah AS, Hoekstra PT, Casacuberta-Partal M, Coffeng LE, Corstjens PLAM, Greco B, et al. Sensitive diagnostic tools and targeted drug administration strategies are needed to eliminate schistosomiasis. Lancet Infect Dis. 2020;20: e165–e172. doi:10.1016/S1473-3099(20)30254-1

37. Anjorin S, Nabatte B, Mpooya S, Tinkitina B, Opio CK, Kabatereine NB, et al. The epidemiology of periportal fibrosis and relevance of current *Schistosoma mansoni* infection: a population-based, cross-sectional study. 2023. doi:10.1101/2023.09.15.23295612

38. Booth M, Vennervald BJ, Kenty L, Butterworth AE, Kariuki HC, Kadzo H, et al. Micro-geographical variation in exposure to *Schistosoma mansoni* and malaria, and exacerbation of splenomegaly in Kenyan school-aged children. BMC Infect Dis. 2004;4: 13. doi:10.1186/1471-2334-4-13

39. Coalition for Operational Research on Neglected Tropical Diseases (COR-NTD). Schistosomiasis Oversampling Study: Survey Strategy Selection Meeting Report. 2023. Available: https://www.cor-ntd.org/resources/schistosomiasis-oversampling-study-survey-strategy-selection-meeting-report

40. Kokaliaris C, Garba A, Matuska M, Bronzan RN, Colley DG, Dorkenoo AM, et al. Effect of preventive chemotherapy with praziquantel on schistosomiasis among school-aged children in sub-Saharan Africa: a spatiotemporal modelling study. Lancet Infect Dis. 2022;22: 136–149. doi:10.1016/S1473-3099(21)00090-6

41. Miguel E, Kremer M. Worms: Identifying Impacts on Education and Health in the Presence of Treatment Externalities. Econometrica. 2004;72: 159–217. doi:10.1111/j.1468-0262.2004.00481.x

42. Seto EYW, Lee YJ, Liang S, Zhong B. Individual and village-level study of water contact patterns and *Schistosoma japonicum* infection in mountainous rural China: S. Japonicum exposure in China. Trop Med Int Health. 2007;12: 1199–1209. doi:10.1111/j.1365-3156.2007.01903.x

43. Bethony J, Williams JT, Brooker S, Gazzinelli A, Gazzinelli MF, LoVerde PT, et al. Exposure to *Schistosoma mansoni* infection in a rural area in Brazil. Part III: household aggregation of water-contact behaviour. Trop Med Int Health. 2004;9: 381–389. doi:10.1111/j.1365-3156.2004.01203.x

44. Seto EYW, Sousa-Figueiredo JC, Betson M, Byalero C, Kabatereine NB, Stothard JR. Patterns of intestinal schistosomiasis among mothers and young children from Lake Albert, Uganda: water contact and social networks inferred from wearable global positioning system dataloggers. Geospatial Health. 2012;7: 1. doi:10.4081/gh.2012.99

45. Phillips AE, Gazzinelli-Guimaraes PH, Aurelio HO, Ferro J, Nala R, Clements M, et al. Assessing the benefits of five years of different approaches to treatment of urogenital schistosomiasis: A SCORE project in Northern Mozambique. Steinmann P, editor. PLoS Negl Trop Dis. 2017;11: e0006061. doi:10.1371/journal.pntd.0006061

46. Musuva R, Odiere M, Mwinzi P, Omondi I, Rawago F, Matendechero S, et al. Unprotected water sources and low latrine coverage are contributing factors to persistent hotspots for schistosomiasis in western Kenya. PLoS ONE. 2021;16. doi:10.1371/journal.pone.0253115

47. Assare R, N’Tamon R, Bellai L, Koffi J, Mathieu T, Ouattara M, et al. Characteristics of persistent hotspots of *Schistosoma mansoni* in western Cote d’Ivoire. Parasit Vectors. 2020;13. doi:10.1186/s13071-020-04188-x

48. Graeff-Teixeira C, Favero V, Pascoal VF, de Souza RP, Rigo F de V, Agnese LHD, et al. Low specificity of point-of-care circulating cathodic antigen (POC CCA) diagnostic test in a non-endemic area for schistosomiasis mansoni in Brazil. Acta Trop. 2021;217: 105863. doi:10.1016/j.actatropica.2021.105863

49. Peralta JM, Cavalcanti MG. Is POC-CCA a truly reliable test for schistosomiasis diagnosis in low endemic areas? The trace results controversy. Hsieh MH, editor. PLoS Negl Trop Dis. 2018;12: e0006813. doi:10.1371/journal.pntd.0006813

50. Pinot de Moira A, Fulford AJC, Kabatereine NB, Ouma JH, Booth M, Dunne DW. Analysis of Complex Patterns of Human Exposure and Immunity to *Schistosomiasis mansoni*: The Influence of Age, Sex, Ethnicity and IgE. PLoS Negl Trop Dis. 2010;4: e820. doi:10.1371/journal.pntd.0000820

51. Masaku J, Njomo DW, Njoka A, Okoyo C, Mutungi FM, Njenga SM. Soil-transmitted helminths and schistosomiasis among pre-school age children in a rural setting of Busia County, Western Kenya: a cross-sectional study of prevalence, and associated exposures. BMC Public Health. 2020;20. doi:10.1186/s12889-020-08485-z

52. Altman DG, Royston P. The cost of dichotomising continuous variables. BMJ. 2006;332: 1080.1. doi:10.1136/bmj.332.7549.1080

53. World Health Organization and UNICEF. Safely managed drinking water - thematic report on drinking water 2017. Geneva: World Health Organization and UNICEF; 2017.

54. World Resources Institute. Waterbodies in Uganda. 2010 [cited 16 Apr 2024]. Available: https://datasets.wri.org/dataset/waterbodies-in-uganda

55. Wood S. mgcv: Mixed GAM Computation Vehicle with Automatic Smoothness Estimation. Available: https://cran.r-project.org/web/packages/mgcv/index.html

56. Clyde M. BAS: Bayesian Variable Selection and Model Averaging using Bayesian Adaptive Sampling. Available: https://cran.r-project.org/web/packages/BAS/index.html

57. Clyde MA, Ghosh J, Littman ML. Bayesian Adaptive Sampling for Variable Selection and Model Averaging. J Comput Graph Stat. 2011;20: 80–101. doi:10.1198/jcgs.2010.09049

58. Liang F, Paulo R, Molina G, Clyde MA, Berger JO. Mixtures of *g* Priors for Bayesian Variable Selection. J Am Stat Assoc. 2008;103: 410–423. doi:10.1198/016214507000001337

59. Bayarri MJ, García-Donato G. Extending conventional priors for testing general hypotheses in linear models. Biometrika. 2007;94: 135–152.

60. Berger JO, Pericchi LR, Ghosh J, Samanta T, De Santis F, Berger J, et al. Objective Bayesian methods for model selection: Introduction and comparison. Lect Notes-Monogr Ser. 2001; 135–207.

61. Barbieri MM, Berger JO. Optimal predictive model selection. Ann Stat. 2004;32. doi:10.1214/009053604000000238

